# Genetic Evolution of Pediatric Acute Myeloid Leukemia and Its Contribution to Disease Relapse

**DOI:** 10.1101/2025.11.04.25339472

**Authors:** Masayuki Umeda, Pandurang Kolekar, Anna LW Huskey, Shruthi Suryaprakash, Jing Ma, Hanxia Li, Tamara Westover, Michael P. Walsh, Guangchun Song, Yanling Liu, Quang Tran, Vidya Balagopal, Lu Wang, Samuel W. Brady, Stan Pounds, Paul E. Mead, Seth E. Karol, Hiroto Inaba, Jeffrey E. Rubnitz, Raul Ribeiro, Xiaotu Ma, Jeffery M. Klco

**Author notes:** Corresponding authors: Jeffery M. Klco and Xiaotu Ma. These authors contributed equally to this work (Masayuki Umeda, Pandurang Kolekar).

## Abstract

Relapse remains the leading cause of mortality in pediatric acute myeloid leukemia (pAML), yet the genetic changes contributing to relapse remain incompletely defined. We performed whole-genome and targeted-capture sequencing of 41 pediatric AML cases with paired relapse samples. Mutational burden increased at relapse, whereas therapy-related mutational signatures were rarely observed and only occasionally associated with pathogenic mutations. Although recurrently enriched mutations at relapse included those in *FLT3*, *WT1*, and *TP53*, relapse-fated subclones were frequently marked only by non-pathogenic or non-coding variants. Longitudinal deep sequencing (∼5000X) coupled with paired serial monitoring by WGS (∼100X) in eight patients showed rapid depletion of major clones after induction therapy, whereas subclones often displayed variable chemosensitivity. Relapse-specific mutations emerged only late, suggesting that selection of pre-existing clones is the predominant mechanism of relapse. Together, our data indicate that pAML relapse arises through the selection of pre-existing clones with diverse trajectories.

## INTRODUCTION

Pediatric acute myeloid leukemia (pAML) accounts for approximately 20% of acute pediatric leukemias and has a long-term survival rate of nearly 70%. Although there have been marginal survival improvements for children with pAML over the past few decades, this is largely due to enhanced supportive care and the intensification of treatments (1, 2). While many patients achieve remission during induction therapies, disease relapse remains common, frequently with aggressive clinical features and high-risk genetics, and is the major cause of mortality. Thus, understanding the biology behind disease relapse will be a fundamental step toward improving clinical outcomes of pAML.

Relapses of acute leukemia can be attributed to multiple mechanisms. Initiating driver alterations are a major determinant of outcomes of pAML, with relapsed disease enriching for specific driver alterations such as *KMT2A* rearrangements (*KMT2A*r), *UBTF* tandem duplications (*UBTF*-TD), or *NUP98* rearrangements (*NUP98*r), which show primary resistance to conventional chemotherapy or high measurable/minimal residual disease (MRD) (3, 4). Also, the enrichment of specific cooperating alterations at relapse, such as *WT1* or *FLT3* mutations, indicates that these somatic alterations render cells with chemo-resistance properties or a growth advantage over cells without mutations (3, 5). However, these alterations are also often found at diagnosis, contrasting to the acquisition of chemotherapy-related/induced somatic mutations during chemotherapy for B-cell acute lymphoblastic leukemia, such as *NT5C2* and *NR3C1* mutations with thiopurine treatment (6) or AML treated with FLT3 inhibitors (7) or menin inhibitors (8, 9). Despite the known genotoxicity of cytotoxic agents used in AML therapies, whether these drugs can promote gene mutations that drive chemoresistance, and ultimately relapse, remains to be studied.

In this study, we profiled 39 diagnosis-relapse (D-R) pairs and two relapse-relapse (R-R) pairs of pAML using whole-genome sequencing (WGS) and target-capture sequencing (TCS), revealing that the majority of pathogenic mutations are identifiable at diagnosis, except for a subset of mutations in *FLT3*, *WT1*, or *TP53.* Comparison of genome-wide genetic profiles at diagnosis and relapse showed the frequent expansion of specific genetic clones at relapse, whereas relapse-fated subclones were not always marked by pathogenic mutations. Longitudinal deep sequencing, supported with ∼100X WGS, of eight cases showed that genetic clones rapidly decline after induction therapy at variable rates, indicating differential chemosensitivity, whereas the emergence of clones with new mutations was a late event, possibly contributing to clonal competition within residual diseases.

## METHODS

### Patient cohort

Samples from patients with relapsed AML were obtained with written informed consent using a protocol approved by the institutional review board (Supplementary Table 1). Samples for WGS (n=39) and TCS (n=97) were newly sequenced in this study, and the rest of the data were obtained from previous publications (Supplementary Tables 1-2).

### Mutational profiling with WGS and TCS

We newly subjected DNA from 16 samples to WGS targeting 40X coverage, followed by mapping to the reference human genome assembly GRCh37-lite with bwa (10). Combined with previously published data, all 41 pairs had WGS data with a median coverage of 49X, which underwent paired analyses with germline samples. Specifically, single-nucleotide variants (SNVs) and insertions/deletions (indels) were called by Bambino (11), copy number variations (CNVs) by CONSERTING (12), and structural variations (SVs) by CREST (13). Detected coding variants were confirmed by manual inspection. The pathogenicity of mutations (pathogenic/likely pathogenic: P/LP) was assessed based on the literature and known function. For longitudinal MRD monitoring, we performed higher-depth WGS for 23 samples from 8 patients, targeting 100X coverage with a median coverage of 104X)

A custom capture panel covering 1.3 Mb genomic regions, including 5979 SNVs/indels and 395 SVs identified in WGS data in this study, was used for validation. Libraries were prepared from 37 patients, with the remaining four patients not having sufficient material for additional sequencing. Pooled samples were sequenced, targeting 500X coverage, and mapped to the GRCh37-lite with bwa (10), with a median coverage of 514X. SNVs were called using SequencErr (14), and indels and SVs were called using SVIndelGenotyper (15). Alterations with supporting reads more than background levels were regarded as real (*P*<0.05 by binomial tests). Remission samples for eight cases were also sequenced using the same pipeline, with a median coverage of 4890X. Alterations with supporting reads more than background levels were regarded as real (*P*<0.05 by binomial tests) as described previously (6). Diagnosis- or relapse-enriched alterations were defined as variants with significant changes in variant allele frequencies (VAF) using WGS data (false discovery rate: FDR-adjusted *P*-value<0.05 with Fisher’s exact test followed by the Benjamini-Hochberg adjustment). Relapse-specific mutations were defined as mutations undetectable at diagnosis with both WGS and TCS, whereas the possibility that mutated clones pre-existed at low frequencies below the detection limit cannot be excluded.

We compared higher-depth WGS (median depth 104X) and deep TCS (median depth 4,890X) platforms for variant clearance using 23 longitudinal samples from eight patients. At each time point, cumulative variant allele frequency (cVAF) was calculated as the ratio of summed mutant reads to summed total reads across all detected somatic markers for each platform. *Overall concordance* between WGS and TCS was assessed using Pearson’s correlation of cVAF values across all 23 samples. Given the limited number of longitudinal samples per patient (1–5 time points; median, 2), patient-wise concordance between platforms was estimated by Pearson’s correlation of VAFs across all valid loci with a minimum depth of 20X.

## RESULTS

### Study cohort of relapsed pAML

We established a cohort of 41 pediatric patients with relapsed AML from clinical trials sponsored by St. Jude Children’s Research Hospital (16–21) (Fig.1A, Supplementary Table 1). The cohort represented major molecular subtypes of pAML (Fig.1B), such as *KMT2A*-rearrangements (*KMT2A*r: n=13, 31.7%) and *NUP98r* (n=6, 14.6%), as well as *NPM1* mutations (n=3, 7.4%), *RUNX1::RUNX1T1* (n=2, 4.9%), or *UBTF* tandem duplications (n=2). Among 39 D-R pairs, the median age at diagnosis was 10.6 years (range: 0.7-20.2), and the days between diagnosis and relapse for D-R pairs were 335 days (175-1773). All diagnosis cases received cytotoxic chemotherapy based on anthracycline, cytarabine, and etoposide (Fig.1C), whereas treatment protocols, which included immunotherapy or molecularly targeted agents, varied according to their assigned clinical trials, such as gemtuzumab ozogamicin: GO in AML02 (17) or sorafenib in AML08 (18). Twelve patients (29.3%) received allogeneic hematopoietic stem cell transplant (allo-HSCT), and seven patients received NK-cell transfusion (17.1%) between two timepoints.

**Figure 1.**
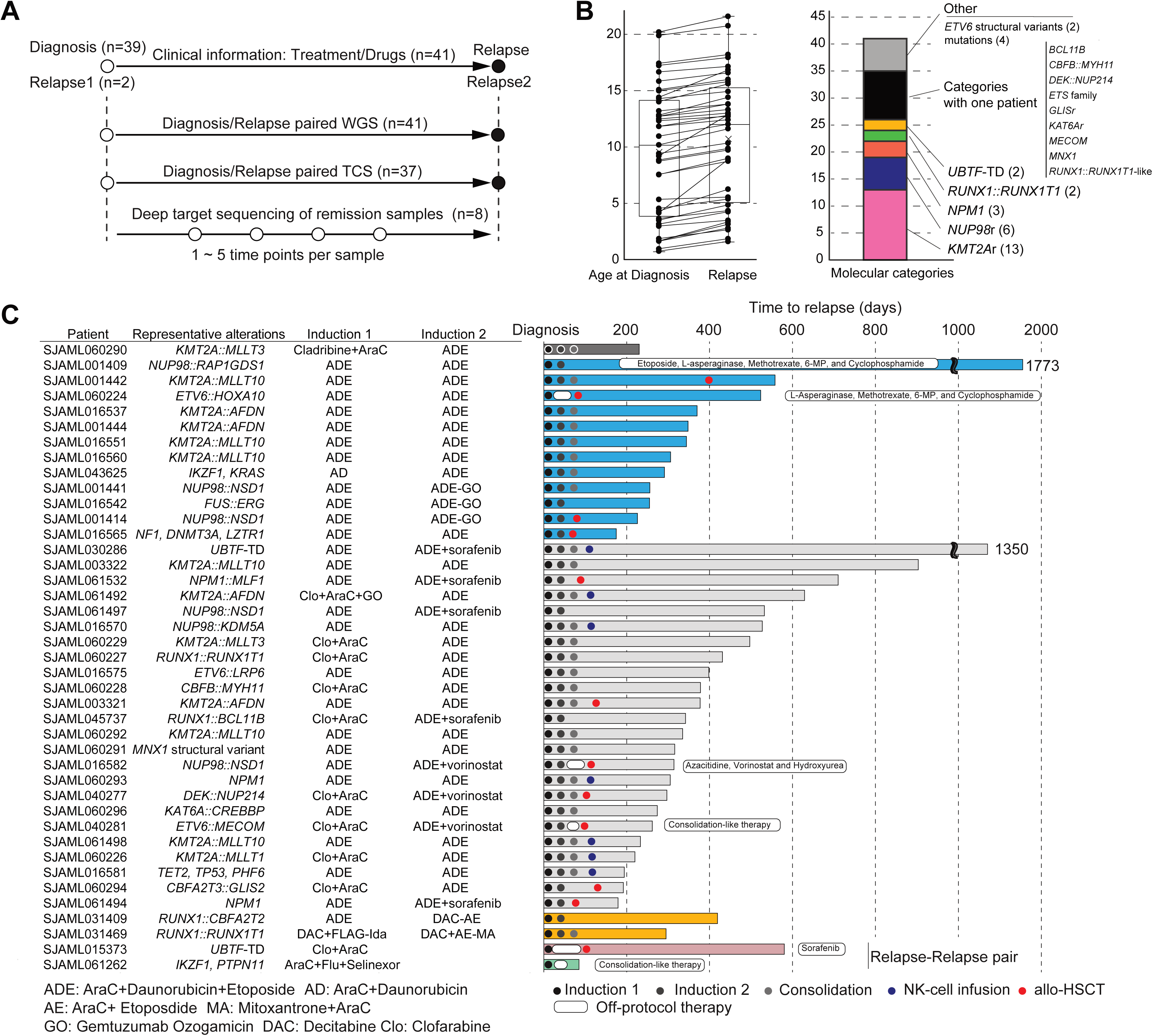
Study cohort of diagnosis-relapse paired pediatric acute myeloid leukemia (pAML). A. Study design showing numbers of patients and timing of sample collection of 41 patient pairs. B. Patient characteristics (age, left; molecular categories, right). C. Clinical characteristics of individual patients within the cohort, including treatments and days to relapse. Lines of the box plots for patient age represent the 25% quantile, median, and 75% quantile. The upper whiskers represent the higher value of maxima or 1.5 x interquartile range (IQR), and the lower whiskers represent the lower value of minima or 1.5 x IQR. Abbreviations. WGS: whole-genome sequencing, TCS: target-capture sequencing, *UBTF*-TD: *UBTF*-tandem duplication, allo-HSCT: allogeneic hematopoietic stem cell transplant.

### Genomic profiling of pAML at diagnosis and relapse

Paired WGS analysis identified 22245 SNVs, 156 indels, 377 CNVs, and 395 SVs (Supplementary Tables 2-6). TCS of selected alterations from 37 patients validated 98.8% (7656/7752) of SNV/indel calls and 95.0% (477/502) SV calls at diagnosis and relapse (Supplementary Figure 1). Among 139 pathogenic or likely pathogenic (P/LP) co-occurring alterations (Fig.2A), *WT1* (n=19) and *NRAS* (n=17) were most commonly mutated in the cohort, followed by *FLT3* (internal tandem duplication-ITD: n=6, tyrosine kinase domain-TKD: n=7, juxtamembrane domain-JMD: n=3) and *SETD2* (n=8).

**Figure 2.**
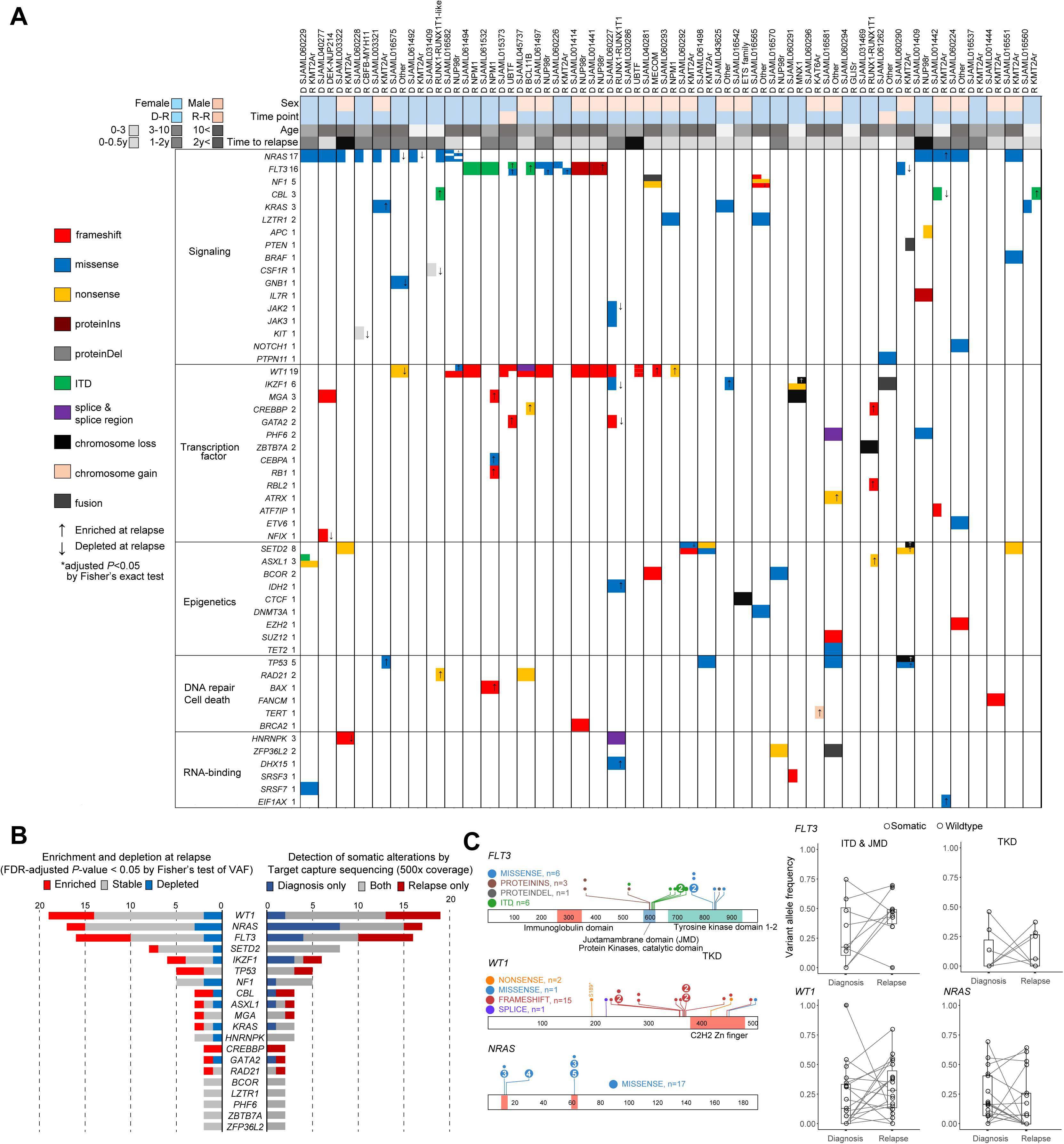
Genomic profiling of pAML at diagnosis and relapse. A. Mutational profiles of pathogenic or likely pathogenic (P/LP) somatic alterations in 39 diagnosis-relapse pairs and two relapse-relapse pairs. Each pair is shown side-by-side. The colors of the heatmap show types of alterations, and arrows indicate enrichment or depletion at relapse. Patient annotations on the top show sex, time points, age, and time to relapse of each patient pair. B. Bar graphs showing numbers of significantly enriched or depleted mutations between two time points of 41 patient pairs (left: red-enrichment, gray-stable, and blue-depletion), and mutations detected at the two time points (right: dark red-relapse only, dark gray-both time points, dark blue-diagnosis only). C. Mutational patterns of *FLT3* (left-top), *WT1* (left-middle), and *NRAS* (left-bottom), and changes of variant allele frequencies between two time points. The colors of dots indicate types of mutations. Lines of the box plots represent the 25% quantile, median, and 75% quantile. The upper and lower whiskers represent the higher value of maxima or 1.5 x IQR and the lower value of minima or 1.5 x IQR, respectively.

The total number of co-occurring P/LP mutations was generally small and did not significantly differ between time points, with a median (range) of 3 (0-9) at diagnosis and 3 (0-6) at relapse (*P*=0.48). Among 139 P/LP cooperating mutations, 38 mutations in 22 genes were enriched (with significantly higher VAF) at relapse. *FLT3* (n=6), *WT1* (n=5), and *TP53* (n=3) were frequently observed (Fig.2B), suggesting that clones harboring these mutations may have a competitive advantage. Notably, 37 alterations, including *FLT3* (n=6), *WT1* (n=6), *TP53*, *CREBBP*, *NRAS*, *IKZF1*, and *CBL* alterations (n=2 each), were detected exclusively at relapse, despite deep TCS profiling (500x) at diagnosis, similar to previous findings (22).

This indicates that these mutations were either present in minor clones below the limit of detection (∼1.0% VAF) at diagnosis or were acquired during or after chemotherapy. Interestingly, clones with these mutations do not always expand at relapse, with *WT1* and *FLT3* alterations showing various patterns of depletion and enrichment (Fig.2C). *NRAS* mutations at diagnosis were frequently decreased to undetectable levels at relapse (7/15: 46.7%), suggesting that clones with *NRAS* mutations are more sensitive to chemotherapy, especially cytarabine (23, 24), or less competitive during disease progression. These data suggest that AML cell populations can acquire somatic mutations during disease progression, whereas clones with pathogenic somatic mutations can selectively expand or decline during disease progression.

### Somatic mutations acquired during disease progression

The mutational burden (SNV/indel) showed a significant increase at relapse compared to diagnosis; median mutations per sample were 216 (range: 10-838) at diagnosis and 376 (71-1893) at relapse (*P*=9.7×10^-6^, Wilcoxon signed-rank test, Fig.3A). SV and CNV were also more frequently observed at relapse (median SV: 4 vs 8, *P*=6.2×10^-5^, median CNV: 3 vs 5, *P*=7.8×10^-3^), although the detection of subclonal CNV (< 20%) can be underestimated by WGS (25). The mutational burden at diagnosis had a strong correlation with age (Spearman’s coefficient: *R*=0.68, *P*=1.8×10^-6^, Fig.3B), whereas one case with a *TP53* mutation (SJAML016581) showed an outlier high mutation burden. Contrarily, mutations newly detected at relapse had only a weak, non-significant correlation (*R*=0.18, *P*=0.28) with time to relapse.

**Figure 3.**
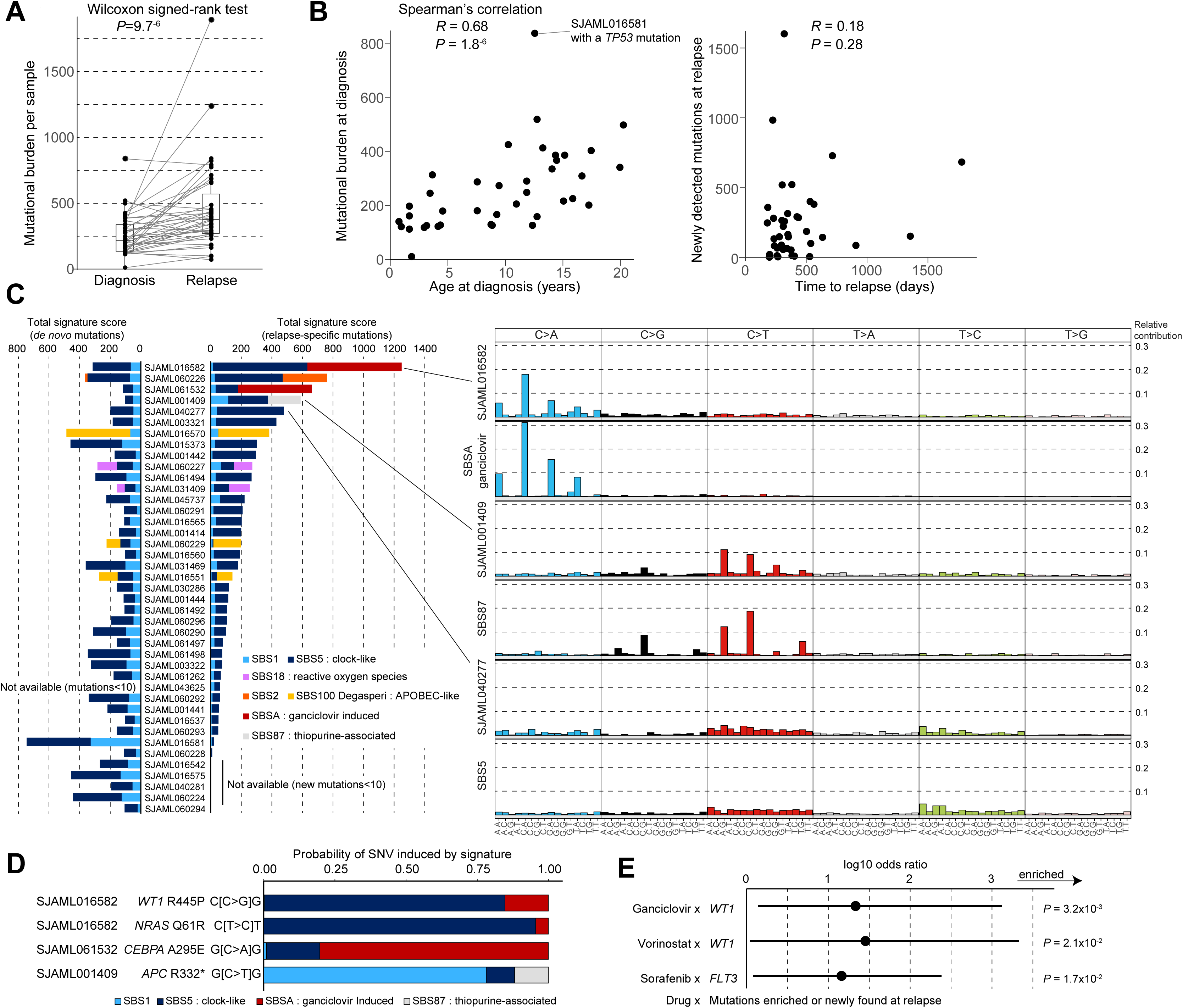
Somatic mutations acquired during disease progression. A. Comparison of mutational burdens at diagnosis and relapse. *P*-value was calculated by the Wilcoxon signed-rank test. B. Scatter plots of age at diagnosis and mutational burden at diagnosis (left) and time to relapse (days) and newly detected mutations at relapse (right). Spearman’s rank correlation was used to assess the coefficient and its statistical significance. C. Bar plots showing the estimated contribution of known mutational signatures to somatic mutation found at diagnosis (left) and newly detected at relapse (middle). The colors of bars indicate the types of known mutational signatures. The right panel shows representative cases compared with known mutational signatures mainly contributing to the case (top-SJAML016582 and SBSA-ganciclovir, mid: SJAML001409 and SBS87 (thiopurine-associated), bottom: SJAML040277 and SBS5-clock-like). D. Contribution of mutational signatures to pathogenic somatic mutations found in patients with drug-associated mutational signatures. E. Odds ratio (dots), 95% confidence interval (lines), and statistical significance between drugs used for treatment and newly detected somatic mutations (*FLT3* and *WT1*) assessed by Fisher’s exact test. Multiple testing adjustment is not applied due to the limited sample size for this explorative analysis.

We then investigated the mutational signatures (26) of somatic mutations found at diagnosis (“de novo mutations”) and those detected only at relapse (“relapse-specific mutations”) to infer the underlying mechanism of mutagenesis (Fig.3C, Supplementary Table 7). De novo mutations were typically enriched for Single Base Substitution (SBS) Signatures SBS1 and SBS5, which are associated with spontaneous mutagenesis consistent with the correlation between mutation number and age at diagnosis. An exception is the presence of SBS18 (reactive oxygen species-related) in two *RUNX1*-fusion cases (27) or SBS2/SBS100 (APOBEC-like) in *KMT2A*r and *NUP98*r cases (n=4). Relapse-specific mutations also exhibited predominantly SBS1/SBS5 signatures coupled with the enrichment of distinct treatment-related signatures in some cases. For example, SBS87 (thiopurine-associated) was found in a patient (SJAML001409) treated with 6-mercaptopurine (6-MP) for off-protocol therapy, and a ganciclovir-related signature was observed in two patients (SJAML016582, SJAML061532) who received ganciclovir for treatment or prophylaxis of cytomegalovirus infection (28). Notably, samples with these treatment-associated signatures harbored P/LP mutations that can be explained by the underlying mutagenic mechanisms (Fig.3D); however, the probabilities were overall low. These data suggest that therapy-related mutagenesis may cause pathogenic mutations in rare cases (eg, *CEBPA* in SJAML016582), but its overall contribution to relapse appears modest, and additional non-genetic mechanisms are likely involved.

We also examined associations between recurrently enriched or acquired P/LP mutations (n=21) as resistance mechanisms for treatment, acknowledging the limited statistical power due to the small sample size for multiple testing adjustment. *FLT3* mutations were associated with sorafenib treatment (*P*=0.017), and three of four acquired *FLT3-*TKD mutations were found in sorafenib-treated patients (*P*=7.1×10^-3^, Fig.3E), consistent with the known association between sorafenib resistance and *FLT3*-TKD mutations (29). We observed alternative *FLT3*-ITDs in two of these patients (*P*=0.018), although they had similar length and insertion sites as the original, differing from *FLT3*-ITDs associated with FLT3 inhibitor resistance (30). As additional findings, *WT1* mutations were enriched in two patients treated with vorinostat (*P*=0.021), which might imply a relationship between *WT1* mutations and vorinostat-induced downregulation of *WT1* expression (31).

### Clonal evolution during disease progression

Given the scarcity of new pathogenic mutations at relapse, we investigated how existing clones contribute to relapse by comparing VAFs of somatic mutations at diagnosis and relapse in each case (n=39, Supplementary Table 8, Supplementary Figure 2A). Clustering of mutations according to VAFs at diagnosis and relapse showed specific patterns in each patient. Initiating and category-defining alterations, such as *RUNX1::RUNX1T1* or *KMT2A*r, were identified with high VAFs in both time points (Fig.4A). However, in most cases (n=37, 94.9%), we observed that mutations in dominant clones or subclones (>10% VAF at diagnosis) were depleted to undetectable levels in the later time points (Fig.4B). Contrarily, alternative subclones marked by somatic mutations with low VAFs at diagnosis became dominant in 34 cases (87.2%). Two relapse-relapse pairs also showed depletion of major clones and expansion of minor clones (Supplementary Figure 2B), indicating that leukemic cells were frequently depleted down to only a few genetic clones during and after treatment, even at the relapse setting.

**Figure 4.**
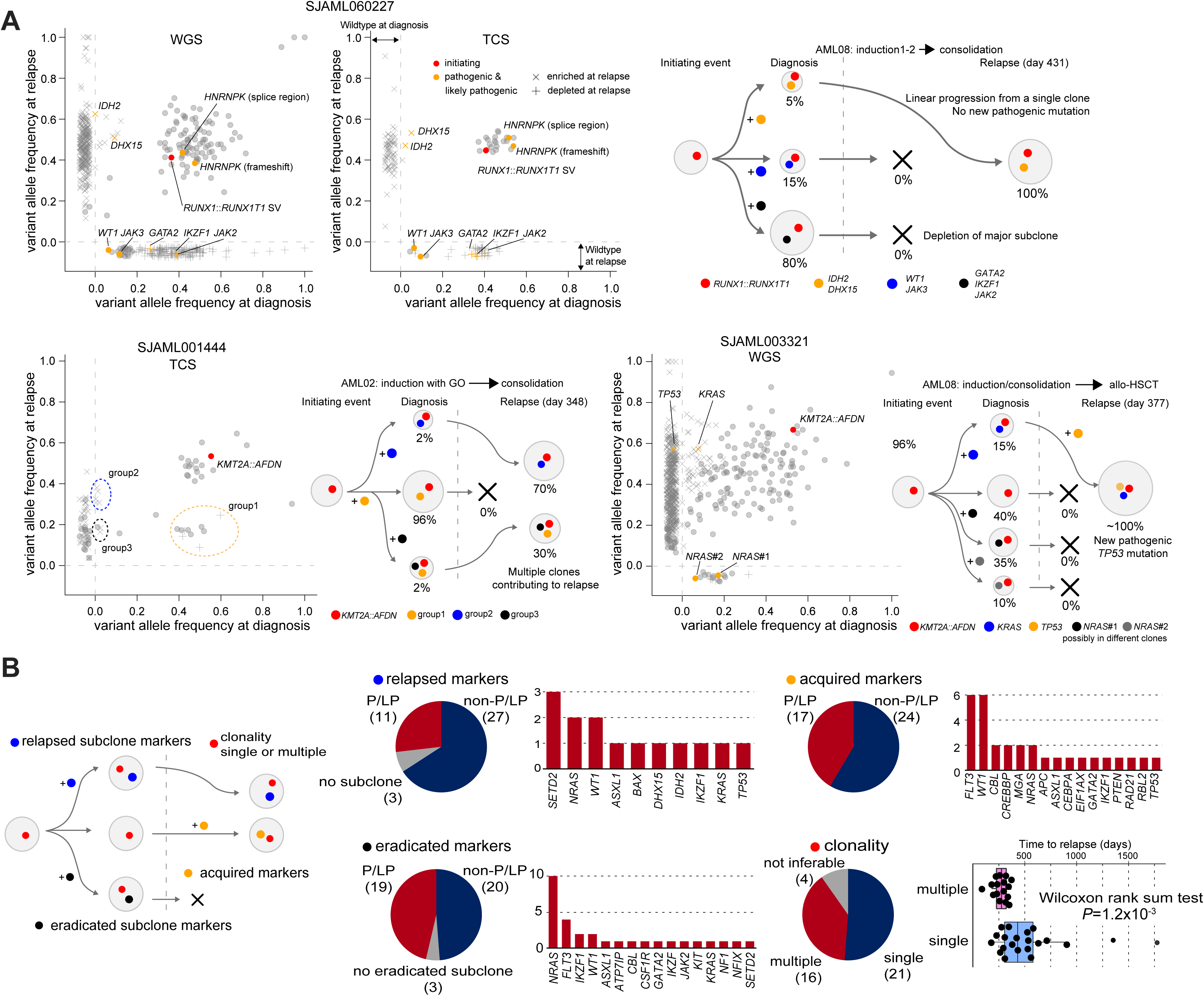
Clonal evolution during disease progression. A. Clonal evolution patterns of three representative cases from the cohort (top: SJAML060277, bottom-left: SJAML001444, and bottom-right: SJAML003321). On 2D plots of variant allele frequencies, the colors of dots show the pathogenicity of somatic alterations, and the shapes of dots show enrichment at each time point. Undetected calls are shown away from the axes. On the schematics of clonal patterns, colored dots show mutations marking clones. B. Schematic (left) and statistical summaries (right) of somatic alterations. Inference of subclones required at least two supporting somatic alterations for each clone. The association with the clonality and time to relapse was assessed with the Wilcoxon rank-sum test. Lines of the box plots represent the 25% quantile, median, and 75% quantile. The upper and lower whiskers represent the higher value of maxima or 1.5 x IQR and the lower value of minima or 1.5 x IQR, respectively.

Among 37 cases with sufficient tumor purity to infer clonality, relapses of 21 cases (56.8%) were inferred to have evolved from single genetic clones (eg, SJAML060227 in Fig.4A), whereas 16 cases (43.2%) were estimated to have a few genetic clones contributing to relapse, with each clone supported by at least two mutations (Fig.4B). Relapses that evolved from multiple clones showed significantly shorter remission (*P*=1.2×10^-3^); multiple: median 266 days (85-370) and single: 547 days (175-1773). These subclones fated for relapse were recurrently marked by *SETD2* (n=3), *WT1* (n=2), or *NRAS* (n=2) mutations, but the majority of cases (27 cases; 73%) had relapses driven by subclones at diagnosis marked by only non-pathogenic or non-coding mutations. Twelve cases acquired P/LP mutations (eg, *WT1* or *FLT3*-TKD, n=3 for each) at relapse, but the remaining 15 cases either had no P/LP cooperating mutations present at diagnosis or acquired at relapse (eg, SJAML001444 in Fig.4A). These data align with experimental models postulating that non-genetic mechanisms, such as epigenetic heterogeneity (32) or stochastic selection, likely confer a competitive advantage to certain clones (33), enabling them to survive therapy and ultimately drive relapse.

### Serial sequencing reveals clonal selection patterns

To gain more insights into how these clones were selected and expanded during therapy, we applied deep sequencing covering somatic mutations identified by WGS (4890X median coverage) on remission samples from eight cases (Supplementary Table 9). Dominant mutations (VAF>40%), including driver alterations, were frequently reduced to 5% or lower after induction, except for one case (SJAML016582) with high VAF (>10%) of dominant mutations during treatment (Fig.5A, Supplementary Figure 3, Supplementary Table 9). Clonal alterations were consistently detected with low VAF (0.1∼10%, two or more mutations in each sample) in the other seven cases throughout the time course, indicating persistent measurable residual disease. Relapse-specific mutations in WGS and TCS data were detected only in samples at the last remission before relapse (by 55-2117 days) in three cases, along with residual dominant mutations. For example, SJAML016582 showed multiple *NRAS*+ subclones at diagnosis, which persisted to day 93, when a new *NRAS* mutation emerged (Fig.5B-C). By relapse at day 310 after allo-HSCT, this clone expanded while another clone obtained a *WT1* mutation, both contributing to relapse. In the remaining five cases, relapse-specific mutations were not consistently detected before clinical relapse, even with deep sequencing (Fig.5E-F, Supplementary Figure 3), suggesting that the acquisition or outgrowth of these mutations is a late event during disease progression.

**Figure 5.**
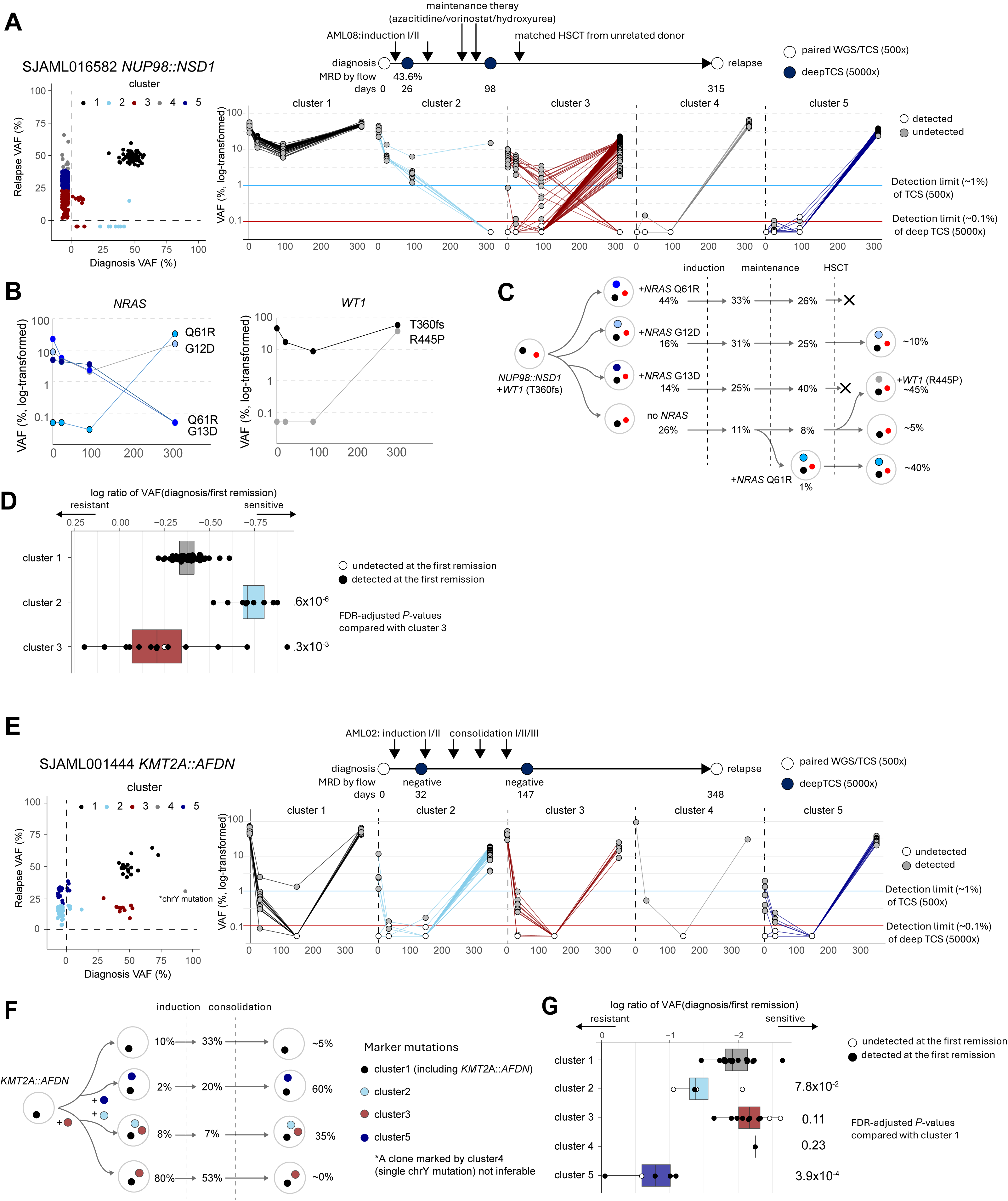
Serial sequencing reveals clonal selection patterns. A. Mutational patterns at diagnosis and relapse of SJAML016582 (left) and clinical courses and changes in VAFs along time points (right). Clusters were defined by hierarchical clustering (Ward method) using VAFs at diagnosis and relapse. B. Changes in VAFs of pathogenic mutations (left: *NRAS* and right: *WT1*). C. Inferred clonal structure from mutational patterns. D. Comparison of log10 fold changes of VAFs of mutations in each cluster at diagnosis and immediately after induction therapy. E. Mutational patterns at diagnosis and relapse of SJAML001444 (left) and clinical courses and changes in VAFs along time points (right). F. Inferred clonal structure from mutational patterns. G. Comparison of log10 fold changes of VAFs of mutations in each cluster at diagnosis and immediately after induction therapy. For plots and fold changes of calls with no mutant read count, a VAF of 0.0005 (half of the expected limit of detection of deep sequencing with 5000x coverage) was used as a value. Statistical significance of the reduction of VAFs was assessed by the Wilcoxon rank-sum test, followed by adjustment by the Benjamini-Hochberg procedure. Lines of the box plots represent the 25% quantile, median, and 75% quantile. The upper and lower whiskers represent the higher value of maxima or 1.5 x IQR and the lower value of minima or 1.5 x IQR, respectively.

We also compared VAFs at diagnosis and immediately after induction therapy in four cases with somatic alterations that remained detectable after therapy, enabling inference of the relative chemosensitivity of each genetic subclone. Compared with dominant mutations, subclonal mutations showed variable, cluster-specific reduction patterns in all cases (adjusted *P*-value<0.05, Fig.5D, Fig.5G, Supplementary Figure 3). Mutations depleted at relapse often showed greater reduction than dominant mutations, while subclones selected at relapse were relatively less sensitive and maintained, despite not necessarily harboring P/LP coding mutations. These data suggest that relapse often originates from subclones that survive chemotherapy, potentially through both genetic and non-genetic mechanisms, with newly acquired cooperating alterations mainly contributing to their competitive advantage later during progression of the disease.

### Longitudinal AML monitoring reveals complementary strengths of WGS and targeted deep sequencing

Monitoring clonal clearance and potentially measurable residual disease with targeted deep sequencing is a powerful, yet potentially limited strategy for pAML due to frequent lack of somatic mutations covered by commercially available sequencing panels that are primarily designed for adult AML. To evaluate the feasibility of genome-wide tracking for clonal clearance, we performed higher-depth WGS (median coverage: 104X) on 23 longitudinal remission samples from eight patients (1–5 time points per patient; median: ∼3) and compared variant detection with matched TCS (median coverage: 4965X; Supplementary Tables 2 and 10). As expected, deep TCS detected more coding variants than high-depth WGS (264 versus 118 among 393 shared somatic markers between diagnosis and relapse, *P*<0.001) in longitudinal samples (Supplementary Figure 4). In contrast, WGS, by definition, enables tracking of a substantially broader repertoire of coding and non-coding mutations, including passenger variants (median 299, range: 121-834 for somatic SNV/indels found at diagnosis for these eight cases; Supplementary Table 6 and Supplementary Figure 5), thereby increasing the number of informative markers without being limited by panel design or requiring patient-specific customization.

We first assessed somatic mutations shared between diagnosis and relapse samples, representing putative ancestral clonal markers for retrospective variant tracking (34–761 markers per patient; median: 277; Fig. 6A and Supplementary Figure 5). We then evaluated diagnosis-restricted somatic mutations alone (121–834 markers; median, 299; Fig. 6B) for prospective variant tracking, representing the actual clinical situation. As observed in the TCS data, comparisons of these markers showed that a large fraction (median: 89%, range: 28-98%) of somatic mutations detected at diagnosis are preserved to relapse (Supplementary Tables 6 and 10; Supplementary Figure 5B). Across patients, VAFs of somatic mutations detected at diagnosis declined markedly after induction therapy and throughout remission, before increasing again at relapse. Clonal trajectories reconstructed from high-depth WGS and deep TCS were broadly concordant, with an overall Pearson’s correlation coefficient (*R*) of 0.96 between cumulative VAFs for both shared and prospective marker sets across 23 timepoints (see Methods) indicating that both approaches effectively capture AML disease dynamics during treatment response and relapse (Supplementary Table 11). Patient-wise concordances at valid loci (see Methods) varied notably (*R* = 0.24 to 0.88; Figs. 6A and 6B), as cases with low disease or mutational burden have lower concordance (Supplementary Table 11). Notably, five of eight patients harbored subtype-defining driver structural variants rather than recurrent coding nucleotide mutations. Among these patients, deep TCS detected structural variants supporting the corresponding fusion events in 63% of longitudinal samples (8/13), compared with 31% for high-depth WGS (Supplementary Table 12). These findings highlight the continued value of targeted deep sequencing if the sensitive detection of specific alterations is needed for monitoring, especially for targeted therapies.

**Figure 6.**
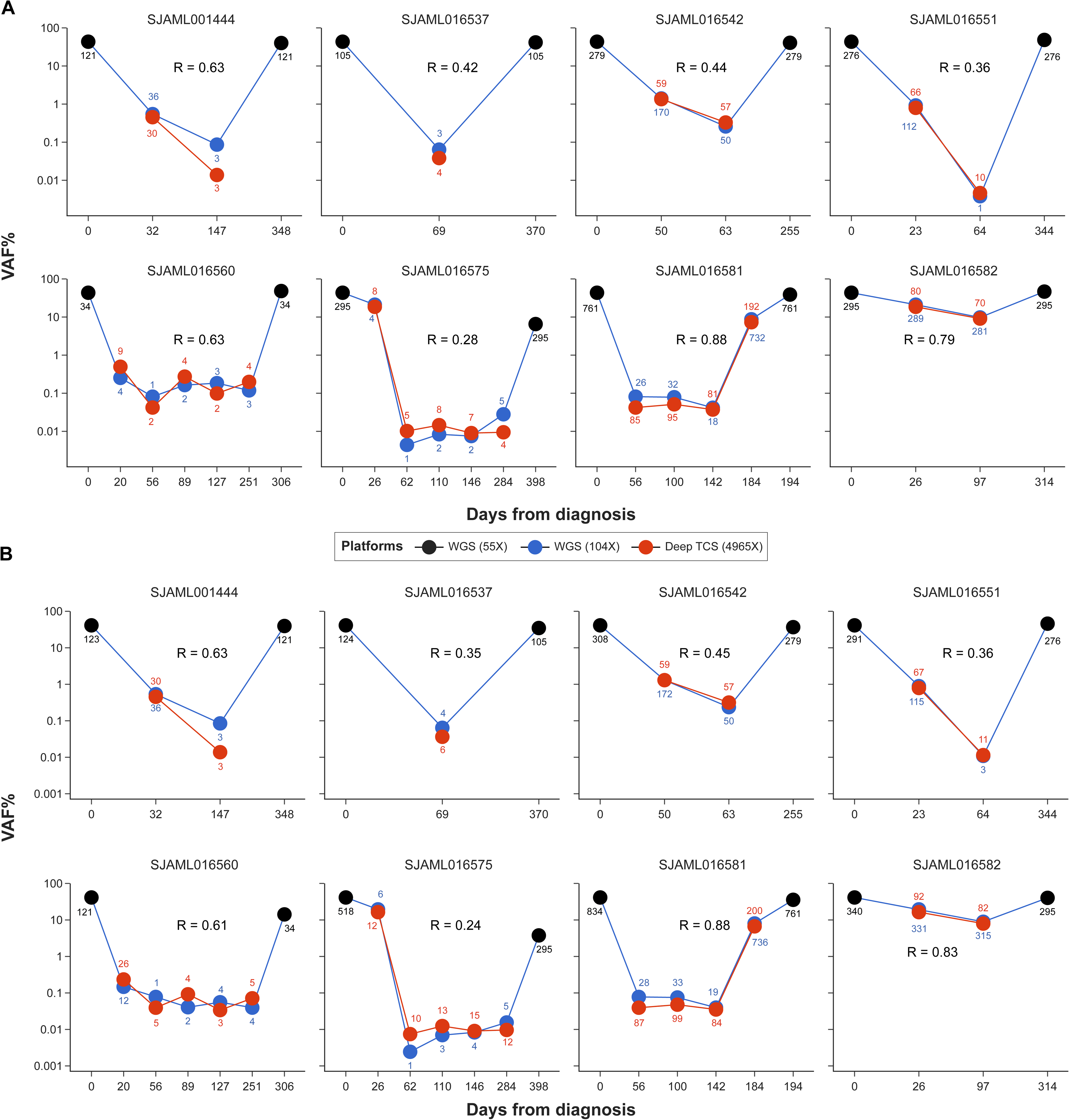
Feasibility of longitudinal WGS sequencing to track the clonal dynamics of pediatric AML. Shown are the cumulative variant allele frequencies (Y-axis) of somatic markers detected using retrospective (panel A) and prospective (panel B) methods across longitudinal samples for eight AML patients using whole-genome sequencing at ∼100X, compared with deep TCS at ∼5000X. The days of sample collection from diagnosis are shown on the X-axis. The points shown in black represent cumulative VAFs of markers detected in diagnosis/relapse timepoint samples sequenced using standard WGS (median depth: 55X), those in blue represent the same for longitudinal samples sequenced using relatively high-depth WGS (median depth: 104X) and those in red represent longitudinal samples with deep TCS data (median depth: 4965X). The numbers of markers detected are shown along points and were used to generate cumulative VAFs from respective sequencing platforms at given sampling points. The patient-wise Pearson’s correlation coefficients (*R*) between VAFs at valid loci (see Methods) captured by high-depth WGS and deep sequencing platforms over longitudinal timepoints are shown.

## DISCUSSION

Relapse remains a major barrier to improving outcomes in pAML. Although initiating alterations at diagnosis are associated with survival, the genome-wide changes between diagnosis and relapse are poorly characterized. To fill the gap, we performed high-resolution mutational profiling of 39 diagnosis– relapse pairs and two relapse-relapse pairs using WGS, complemented by TCS and inferred clonal architecture.

Despite frequent mutations in *WT1* and *SETD2* in this cohort, these alterations were often detectable in subclones at diagnosis. Whereas we expectedly observed a significant increase in mutational burden at relapse, most of these newly acquired mutations were non-pathogenic or located in non-coding regions, with exceptions of a subset of *FLT3* and *WT1* mutations. We also observed a known genetic resistance to sorafenib mediated by *FLT3*-TKD mutations, which supports clinical trials for pediatric AML testing other *FLT3* inhibitors effective for both *FLT3*-ITD and TKD, whereas the other associations require validation in a larger cohort due to the limitation of statistical power of this study.

We next examined the underlying biology of somatic mutations by mutational signature profiling, which showed that the majority of the newly acquired mutations were attributable to spontaneous mutagenesis (SBS1/SBS5). Whereas some cases treated with 6-MP or ganciclovir had mutational signatures associated with treatment, these rarely contributed to the acquisition of pathogenic mutations. Given that 6-MP is not routinely used in standard AML therapy and ganciclovir is being replaced by valganciclovir or letermovir for cytomegalovirus prevention and treatment, our data suggest that therapy-related mutagenesis may not be a primary driver of pAML progression. This pattern contrasts with pediatric ALL, where acquisition of mutations linked to specific drugs is commonly observed (6).

Further comparisons of mutational profiles across patients revealed a common pattern across different treatment contexts; dominant clones at diagnosis were depleted, while alternative minor subclones marked by somatic alterations expanded toward relapse. These alterations were often non-pathogenic or non-coding mutations. Longitudinal deep-targeted sequencing of remission samples in eight patients further showed that genetic subclones differ in their sensitivity to induction therapy and often persist throughout the clinical course. Expansion of clones with newly acquired mutations was observed only at later stages. These observations suggest that relapse frequently arises through the non-deterministic survival of subclones or epigenetic mechanisms, rather than through the persistence of a single population defined by pathogenic mutations.

Genetic clearance (34, 35) or detection of MRD after induction therapies (36) are strong predictors of AML relapse and overall response to therapy. Although not designed or powered to address the predictive power of molecular measurable residual disease determination, our study provides an important direct comparison of targeted deep sequencing to unbiased deep (∼100X) WGS. This is especially relevant as pediatric AMLs often show low mutation burden at diagnosis, and are frequently driven by structural variants, resulting in many commercially available fixed panel approaches being underpowered for pediatric AML. Our deep (100X) WGS method showed the feasibility of prospective residual tumor tracking for pediatric AML, where WGS-based VAF estimates demonstrated sufficient concordance with those estimates established by targeted sequencing. However, the number of detectable variants varied widely across patients and disease stages, ranging from only a few mutations during deep remission to hundreds at diagnosis and relapse. By enabling simultaneous assessment of genome-wide coding and non-coding variants, moderately deep WGS may improve the robustness of MRD monitoring by increasing marker availability, particularly in samples with low disease burden. In such settings, however, rigorous error modeling and careful interpretation of low-frequency variants remain essential. A similar approach was recently described by Shiloh et al, in which WGS was used to follow “leukemia-specific passenger variants”, highlighting the potential clinical value of this approach for monitoring disease burden (37). Despite the differences in genetic makeup for AML across the age spectrum, a WGS strategy for variant monitoring has also shown promise for adult AML. Patient-specific panel/primer design is an alternative, and likely the superior option if the detection of a specific alteration is needed (38, 39). However, this requires the generation and validation of unique clinical reagents for each patient, which remains a challenge for most academic centers. In the future, MRD assessment of pediatric AML with a WGS strategy could be clinically feasible as sequencing costs decrease with technological improvements along with bioinformatic advances.

In addition to the genetic heterogeneity of relapse that we highlighted in this study, transcriptional and epigenetic changes are also likely to shape disease progression and relapse. For example, some studies have demonstrated an enrichment of immature populations at relapse, which has been proposed as a common feature in major subtypes of pAML (40). Further, treatment-specific selective pressures introduced by targeted therapies such as BCL2 inhibitors (41, 42) or menin inhibitors (8, 9) are also likely to shape relapse trajectories. Whereas our study focused on genetic biomarkers of disease to assess therapeutic response and progression, future studies integrating genetic factors with non-genetic and cell state features are warranted to fully understand the mechanisms and trajectory pediatric AML relapses. Recent adult AML-focused studies observed additional layers of genetic complexity using chromatin accessibility patterns that linked genetics with gene expression and differentiation state (32), and these patterns are likely to drive outcome and evolve during relapse. Also, leveraging an improved understanding of the biological mechanisms underlying disease relapse to optimize treatment strategies that address both genetic and epigenetic vulnerabilities will be key to achieving durable cures for children with AML.

## ACKNOWLEDGEMENTS

We thank all the patients and their families for their contribution of the biological specimens used in this study. We also thank the Biorepository, the Flow Cytometry and Cell Sorting Core, and the Hartwell Center for Bioinformatics and Biotechnology at SJCRH for their essential services. The authors thank Ben Huang, MD (UCSF), for a critical review of this manuscript.

## AUTHOR CONTRIBUTIONS

J.M.K and X. M conceptualized the study. J.M.K, M.U, P.K, A.L.W.H, S.S, H.L, M.P.W, G.S, J.M, T.W, Y.L, Q.T, P.M, and X.M collected data. J.M.K, M.U, P.K, H.L, M.P.W, G.S, J.M, T.W, and X.M curated data. M.U, and J.M conducted formal analyses. M.U, J.M, H.L, and X.M performed statistical tests. M.U and P.K made figures. S.K, H.I, J.E.R, and R.R provided resources. M.U and J.M.K wrote the original draft, and all the co-authors reviewed it and were involved in the revision process.

## SUPPLEMENTARY METHOD

### Subject cohorts and sample details

No patient received compensation for enrollment in this study. Patient names were de-identified and replaced with unique research IDs, which are maintained using an honest broker, in accordance with institutional protocols and ethical guidelines. This unique research ID cannot be linked to patient identification outside of the research team. Samples for whole genome sequencing (WGS: n=39), and target capture sequencing (TCS: n=97) were newly sequenced in this study, and the rest of the data were obtained from previous publications (3, 43, 44), European Genome-Phenome Archive under accession number <u>EGAS00001004701</u> (45), or <u>St. Jude Cloud</u> (43) (see details in **DATA AVAILABILITY** and Supplementary Table 1-2).

### Sample processing, library preparation, and sequencing

For newly sequenced samples with insufficient tumor purity (*e.g.*, below 60%), we enriched the leukemic cell population either by flow cytometric sorting or T cell depletion by magnetic beads (EasySep Human CD3 Positive Selection Kit II, StemCell Technologies, Vancouver, BC, Canada, 17851) as we have previously reported(36). CD45^dim^CD33^dim^ positive population was sorted using anti-CD45 PerCP-Cyanine5.5 (eBioscience, San Diego, CA, US, cat# 8045-9459-120, Clone:2D1, RRID: AB_1907397, 1:20 dilution), anti-CD33 APC (eBioscience, cat# 17-0338-42,clone:WM53, RRID:AB_10667893, 1:20 dilution), and DAPI (BD Biosciences, San Jose, CA, US, cat# 564907, RRID:AB_2869624) using FACSAria III instrument and FACS Diva v9.0 (both BD Biosciences) as we have implemented(36). CD34 gating using anti-CD34 PE (Beckman Coulter, Brea, CA, US, cat# IM1459U, Clones:QBEnd10; Immu133; Immu409, RRID:AB_131210, 1:5 dilution) was included depending on each patient sample. Enrichment of the tumor population was confirmed by flow cytometric analysis of the post-sorting samples, generally achieving 90%. Libraries were constructed using the TruSeq DNA PCR-Free Library Prep Kit (Illumina, 20015963) for WGS according to the manufacturer’s instructions. TWIST Biosciences (South San Francisco, CA, US) NGS Target Enrichment Library Preparations with enzymatic fragmentation was used for TCS according to the manufacturer’s protocol. After library quality and quantity assessment, WGS samples were sequenced on HiSeq2000 or 2500 (Illumina, RRID:SCR_020132, RRID:SCR_016383) instruments with paired-end (2 × 101 bp, 2 x 126 bp, or 2 x 151 bp) sequencing using TruSeq SBS Kit v3-HS (Illumina, FC-401-3001) or TruSeq Rapid SBS Kit (Illumina, FC-402-4023) and HiSeq Control Software of the latest version at the time of sequencing. Pooled TCS samples were sequenced using Illumina Novaseq SP 300 cycle kit (paired-end, 150 bp) targeting 500X coverages or Illumina NovaSeq S1 200-cycle kit targeting 5000X for remission samples.

### DNA data mapping and detection of alterations

Both WGS and TCS sequencing data were mapped to the reference human genome assembly GRCh37-lite with bwa (10) (WGS: v0.7.15-r1140 and v0.5.9-r26-dev, RRID:SCR_010910). For WGS, candidate SNVs and Indels were called by Bambino (11) (no version) and classified for putative pathogenicity with PeCanPie/MedalCeremony(46). SV were analyzed using CREST (13) (v1.0), and CNVs were analyzed using CONSERTING (12) (no version). SNVs were called using SequencErr (14) (v2.09), and Indels and SVs were called using SVIndelGenotyper (15, 47) (v1.0, https://github.com/stjude/SVIndelGenotyper). Mutations in relapse samples suspected to be contamination of normal bone marrow cells from allogenic hematopoietic stem cell transplant (allo-HSCT) graft were removed manually.

### Verification of somatic alterations detected by capture sequencing data

To verify somatic mutations using capture sequencing data, we first constructed a background error profile for each marker, described previously as “rotating control” (6). Briefly, for a given marker, the background was established by genotyping the corresponding position across the germline samples of all patients excluding the index patient(s) in whom the marker was initially identified. This yielded a reference set of mutant (nM) and total (nT) read counts representing background noise. The observed nM and nT values for the marker in the index sample were then compared against this background using a binomial test to calculate a p-value. Multiple testing correction was performed using the false discovery rate (FDR) method in Python module “statsmodels”. Markers with a q-value <0.05 were classified as somatic mutations, whereas markers with a q-value ≥ 0.05 were considered wild-type.

### Mutation Signature Analysis

Mutational signature analyses on WGS tier 1-3 SNV calls performed by MutationalPatterns (26) (v3.6.0) on samples with 10 or more SNVs. Mutations found at diagnosis and mutations specific to relapse were analyzed separately to dissect *de novo* somatic mutations and mutations possibly acquired after treatment. *De novo* mutational signature extraction using NMF (non-negative matrix factorization) was initially performed for each WGS sample, followed by testing for the presence of 238 publicly available signatures (COSMIC (48): n=60, Signal (49): n=174, the experimental thiopurine signatures (6) (n=2), the thio_dMMR signature (50), and the ganciclovir signature(28): each n=1). Based on these data, the finalized WGS signature data were obtained by testing for the presence of only the following signatures in each sample: COSMIC signatures 1 and 5 (clock-like), COSMIC signature 2 (APOBEC), COSMIC signature 87 (Thiopurine), COSMIC signature 18 (reactive oxygen species), the ganciclovir signature (28), and the published SBS100 (49). A required cosine similarity increase of 0.03 or more was used for a new signature to be considered detected in a single sample. The probability that individual SNVs were caused by a signature was calculated as carried out in Li et al (6).

### Statistics

One-sided binomial tests for mutation calls were performed using the binom.test function from the stats library (SCR_025968) on the R statistical environment using variant and total allele counts from tumor samples and germline controls as input. Two-sided Fisher’s tests for the enrichment or depletion of somatic mutations were performed with the fisher.test from the stats library. Spearman correlations among clinical parameters and mutational burden were calculated with the cor.test function from the stats library, assuming non-normal distribution and non-linear correlation. Wilcoxon rank sum test for unpaired samples and signed rank test for paired samples were performed using the wilcox.test function from the stat library for comparison of data with non-normal distributions. Kruskal-Wallis test (the kruskal.test function from the stat library) followed by Dunn’s test (the dunn.test library) for multiple comparison of data with non-normal distribution. Benjamini-Hochberg procedure was performed using the p.adjust function when the adjustment for multiple testing was appropriate.

## DATA AVAILABILITY

Genomic analyses in this study were based on the GENCODE GRCh37/hg19, and gnomAD (version 2.1.1, RRID: SCR_014964) was used for classification of germline and somatic mutations. The sequencing data from patient samples newly generated in this study (WGS: n=39, TCS: n=97) have been deposited in the European Genome-Phenome Archive (EGA, RRID:SCR_004944), which is hosted by the European Bioinformatics Institute (EBI), under accession <u>EGAS00001005760</u>. Of the remaining WGS data for 66 samples, 56 samples were sequenced at St. Jude and are available on either EGA or St. Jude Cloud (3, 43, 44), and for the other 10 published cases (45), we downloaded the BAM files from EGA (<u>EGAS00001004701</u>).

Other data generated in this study are available in Supplemental tables or upon request to the corresponding author. We did not use any custom code in this study.

## REFERENCES

1. Bolouri H, Farrar JE, Triche T, Jr., Ries RE, Lim EL, Alonzo TA, et al. The molecular landscape of pediatric acute myeloid leukemia reveals recurrent structural alterations and age-specific mutational interactions. Nat Med. 2018;24(1):103–12.

2. Rubnitz JE, Kaspers GJL. How I treat pediatric acute myeloid leukemia. Blood. 2021;138(12):1009–18.

3. Umeda M, Ma J, Huang BJ, Hagiwara K, Westover T, Abdelhamed S, et al. Integrated Genomic Analysis Identifies UBTF Tandem Duplications as a Recurrent Lesion in Pediatric Acute Myeloid Leukemia. Blood Cancer Discov. 2022;3(3):194–207.

4. Umeda M, Ma J, Westover T, Ni Y, Song G, Maciaszek JL, et al. A new genomic framework to categorize pediatric acute myeloid leukemia. Nat Genet. 2024.

5. Stratmann S, Yones SA, Mayrhofer M, Norgren N, Skaftason A, Sun J, et al. Genomic characterization of relapsed acute myeloid leukemia reveals novel putative therapeutic targets. Blood Adv. 2021;5(3):900–12.

6. Li B, Brady SW, Ma X, Shen S, Zhang Y, Li Y, et al. Therapy-induced mutations drive the genomic landscape of relapsed acute lymphoblastic leukemia. Blood. 2020;135(1):41–55.

7. Kiyoi H, Kawashima N, Ishikawa Y. FLT3 mutations in acute myeloid leukemia: Therapeutic paradigm beyond inhibitor development. Cancer Sci. 2020;111(2):312–22.

8. Perner F, Stein EM, Wenge DV, Singh S, Kim J, Apazidis A, et al. MEN1 mutations mediate clinical resistance to menin inhibition. Nature. 2023;615(7954):913–9.

9. Tiong IS, Ritchie DS, Blombery P. Response and Resistance to Menin Inhibitor in UBTF-Tandem Duplication AML. N Engl J Med. 2024;390(24):2323–5.

10. Li H, Durbin R. Fast and accurate short read alignment with Burrows-Wheeler transform. Bioinformatics. 2009;25(14):1754–60.

11. Edmonson MN, Zhang J, Yan C, Finney RP, Meerzaman DM, Buetow KH. Bambino: a variant detector and alignment viewer for next-generation sequencing data in the SAM/BAM format. Bioinformatics. 2011;27(6):865–6.

12. Chen X, Gupta P, Wang J, Nakitandwe J, Roberts K, Dalton JD, et al. CONSERTING: integrating copy-number analysis with structural-variation detection. Nat Methods. 2015;12(6):527–30.

13. Wang J, Mullighan CG, Easton J, Roberts S, Heatley SL, Ma J, et al. CREST maps somatic structural variation in cancer genomes with base-pair resolution. Nat Methods. 2011;8(8):652–4.

14. Davis EM, Sun Y, Liu Y, Kolekar P, Shao Y, Szlachta K, et al. SequencErr: measuring and suppressing sequencer errors in next-generation sequencing data. Genome Biol. 2021;22(1):37.

15. Kolekar P, Balagopal V, Dong L, Liu Y, Foy S, Tran Q, et al. SJPedPanel: A Pan-Cancer Gene Panel for Childhood Malignancies to Enhance Cancer Monitoring and Early Detection. Clin Cancer Res. 2024;30(18):4100–14.

16. Rubnitz JE, Crews KR, Pounds S, Yang S, Campana D, Gandhi VV, et al. Combination of cladribine and cytarabine is effective for childhood acute myeloid leukemia: results of the St Jude AML97 trial. Leukemia. 2009;23(8):1410–6.

17. Rubnitz JE, Inaba H, Dahl G, Ribeiro RC, Bowman WP, Taub J, et al. Minimal residual disease-directed therapy for childhood acute myeloid leukaemia: results of the AML02 multicentre trial. Lancet Oncol. 2010;11(6):543–52.

18. Rubnitz JE, Lacayo NJ, Inaba H, Heym K, Ribeiro RC, Taub J, et al. Clofarabine Can Replace Anthracyclines and Etoposide in Remission Induction Therapy for Childhood Acute Myeloid Leukemia: The AML08 Multicenter, Randomized Phase III Trial. J Clin Oncol. 2019;37(23):2072–81.

19. Alexander TB, Lacayo NJ, Choi JK, Ribeiro RC, Pui CH, Rubnitz JE. Phase I Study of Selinexor, a Selective Inhibitor of Nuclear Export, in Combination With Fludarabine and Cytarabine, in Pediatric Relapsed or Refractory Acute Leukemia. J Clin Oncol. 2016;34(34):4094–101.

20. Karol SE, Alexander TB, Budhraja A, Pounds SB, Canavera K, Wang L, et al. Venetoclax in combination with cytarabine with or without idarubicin in children with relapsed or refractory acute myeloid leukaemia: a phase 1, dose-escalation study. Lancet Oncol. 2020;21(4):551–60.

21. Inaba H, Rubnitz JE, Coustan-Smith E, Li L, Furmanski BD, Mascara GP, et al. Phase I pharmacokinetic and pharmacodynamic study of the multikinase inhibitor sorafenib in combination with clofarabine and cytarabine in pediatric relapsed/refractory leukemia. J Clin Oncol. 2011;29(24):3293–300.

22. Farrar JE, Schuback HL, Ries RE, Wai D, Hampton OA, Trevino LR, et al. Genomic Profiling of Pediatric Acute Myeloid Leukemia Reveals a Changing Mutational Landscape from Disease Diagnosis to Relapse. Cancer Res. 2016;76(8):2197–205.

23. Neubauer A, Maharry K, Mrozek K, Thiede C, Marcucci G, Paschka P, et al. Patients with acute myeloid leukemia and RAS mutations benefit most from postremission high-dose cytarabine: a Cancer and Leukemia Group B study. J Clin Oncol. 2008;26(28):4603–9.

24. Brendel C, Teichler S, Millahn A, Stiewe T, Krause M, Stabla K, et al. Oncogenic NRAS Primes Primary Acute Myeloid Leukemia Cells for Differentiation. PLoS One. 2015;10(4):e0123181.

25. Wang L, Voss R, Pastor V, Cardenas M, Kumar P, Maciaszek J, et al. Integrated Whole Genome and Transcriptome Sequencing as a Framework for Pediatric and Adolescent AML Diagnosis and Risk Assessment. Res Sq. 2025.

26. Manders F, Brandsma AM, de Kanter J, Verheul M, Oka R, van Roosmalen MJ, et al. MutationalPatterns: the one stop shop for the analysis of mutational processes. BMC Genomics. 2022;23(1):134.

27. Mandell JD, Fisk JN, Cyrenne E, Xu ML, Cannataro VL, Townsend JP. Not only mutations but also tumorigenesis can be substantially attributed to DNA damage from reactive oxygen species in RUNX1::RUNX1T1-fusion-positive acute myeloid leukemia. Leukemia. 2022;36(12):2931–3.

28. de Kanter JK, Peci F, Bertrums E, Rosendahl Huber A, van Leeuwen A, van Roosmalen MJ, et al. Antiviral treatment causes a unique mutational signature in cancers of transplantation recipients. Cell Stem Cell. 2021;28(10):1726–39 e6.

29. Man CH, Fung TK, Ho C, Han HH, Chow HC, Ma AC, et al. Sorafenib treatment of FLT3-ITD(+) acute myeloid leukemia: favorable initial outcome and mechanisms of subsequent nonresponsiveness associated with the emergence of a D835 mutation. Blood. 2012;119(22):5133–43.

30. Kayser S, Schlenk RF, Londono MC, Breitenbuecher F, Wittke K, Du J, et al. Insertion of FLT3 internal tandem duplication in the tyrosine kinase domain-1 is associated with resistance to chemotherapy and inferior outcome. Blood. 2009;114(12):2386–92.

31. Galimberti S, Canestraro M, Khan R, Buda G, Orciuolo E, Guerrini F, et al. Vorinostat and bortezomib significantly inhibit WT1 gene expression in MO7-e and P39 cell lines. Leukemia. 2008;22(3):628–31.

32. Ochi Y, Liew-Littorin M, Nannya Y, Bengtzen S, Piauger B, Deneberg S, et al. Chromatin landscape and epigenetic heterogeneity of acute myeloid leukaemia. Nature. 2026.

33. Fennell KA, Vassiliadis D, Lam EYN, Martelotto LG, Balic JJ, Hollizeck S, et al. Non-genetic determinants of malignant clonal fitness at single-cell resolution. Nature. 2022;601(7891):125–31.

34. Klco JM, Miller CA, Griffith M, Petti A, Spencer DH, Ketkar-Kulkarni S, et al. Association Between Mutation Clearance After Induction Therapy and Outcomes in Acute Myeloid Leukemia. JAMA. 2015;314(8):811–22.

35. Jacoby MA, Spencer DH, Gao F, Uy GL, Burton T, Heath SE, et al. Consolidation Therapy Based on Mutation Clearance in Acute Myeloid Leukemia. NEJM Evid. 2026;5(9):EVIDoa2500352.

36. Umeda M, Ma J, Westover T, Ni Y, Song G, Maciaszek JL, et al. A new genomic framework to categorize pediatric acute myeloid leukemia. Nat Genet. 2024;56(2):281–93.

37. Shiloh R, Dayan D, Geron I, Cohen M, Feuerstein T, Cano-Nistal J, et al. Passenger variants as pan-clonal markers of pediatric AML: Implications for molecular MRD. Hemasphere. 2026;10(8):e70424.

38. Yu K, Dillon LW, Tettero JM, Gui G, Al-Ali RW, Grunwald MR, et al. Genomics of Acute Myeloid Leukemia at Diagnosis and Remission. medRxiv. 2025.

39. Ravindra N, Lack J, Dalgard CL, vanCollenburg E, Corner A, Beppu L, et al. Whole Genome Sequencing Informed Patient Personalized Measurable Residual Disease Assays for Acute Myeloid Leukemia. medRxiv. 2026.

40. Lambo S, Trinh DL, Ries RE, Jin D, Setiadi A, Ng M, et al. A longitudinal single-cell atlas of treatment response in pediatric AML. Cancer Cell. 2023;41(12):2117–35 e12.

41. Pei S, Pollyea DA, Gustafson A, Stevens BM, Minhajuddin M, Fu R, et al. Monocytic Subclones Confer Resistance to Venetoclax-Based Therapy in Patients with Acute Myeloid Leukemia. Cancer Discov. 2020;10(4):536–51.

42. Kuusanmaki H, Dufva O, Vaha-Koskela M, Leppa AM, Huuhtanen J, Vanttinen I, et al. Erythroid/megakaryocytic differentiation confers BCL-XL dependency and venetoclax resistance in acute myeloid leukemia. Blood. 2023;141(13):1610–25.

43. Newman S, Nakitandwe J, Kesserwan CA, Azzato EM, Wheeler DA, Rusch M, et al. Genomes for Kids: The scope of pathogenic mutations in pediatric cancer revealed by comprehensive DNA and RNA sequencing. Cancer Discov. 2021.

44. Umeda M, Ma J, Westover T, Ni Y, Song G, Maciaszek J, et al. Proposal of a new genomic framework for categorization of pediatric acute myeloid leukemia associated with prognosis. Research Square; 2023.

45. Fornerod M, Ma J, Noort S, Liu Y, Walsh MP, Shi L, et al. Integrative Genomic Analysis of Pediatric Myeloid-Related Acute Leukemias Identifies Novel Subtypes and Prognostic Indicators. Blood Cancer Discov. 2021;2(6):586–99.

46. Edmonson MN, Patel AN, Hedges DJ, Wang Z, Rampersaud E, Kesserwan CA, et al. Pediatric Cancer Variant Pathogenicity Information Exchange (PeCanPIE): a cloud-based platform for curating and classifying germline variants. Genome Res. 2019;29(9):1555–65.

47. Shao Y, Tran Q, Kolekar P, Liu Y, Liang Z, Fan L, et al. Analysis of Error Profiles of Indels and Structural Variants in Deep Sequencing Data. SSRN Electronic Journal. 2025.

48. Alexandrov LB, Kim J, Haradhvala NJ, Huang MN, Tian Ng AW, Wu Y, et al. The repertoire of mutational signatures in human cancer. Nature. 2020;578(7793):94–101.

49. Degasperi A, Zou X, Amarante TD, Martinez-Martinez A, Koh GCC, Dias JML, et al. Substitution mutational signatures in whole-genome-sequenced cancers in the UK population. Science. 2022;376(6591).

50. Yang F, Brady SW, Tang C, Sun H, Du L, Barz MJ, et al. Chemotherapy and mismatch repair deficiency cooperate to fuel TP53 mutagenesis and ALL relapse. Nat Cancer. 2021;2(8):819–34.

