## Supplementary material for "Genetic Evolution of Pediatric Acute Myeloid Leukemia and Its Contribution to Disease Relapse": Fig S1

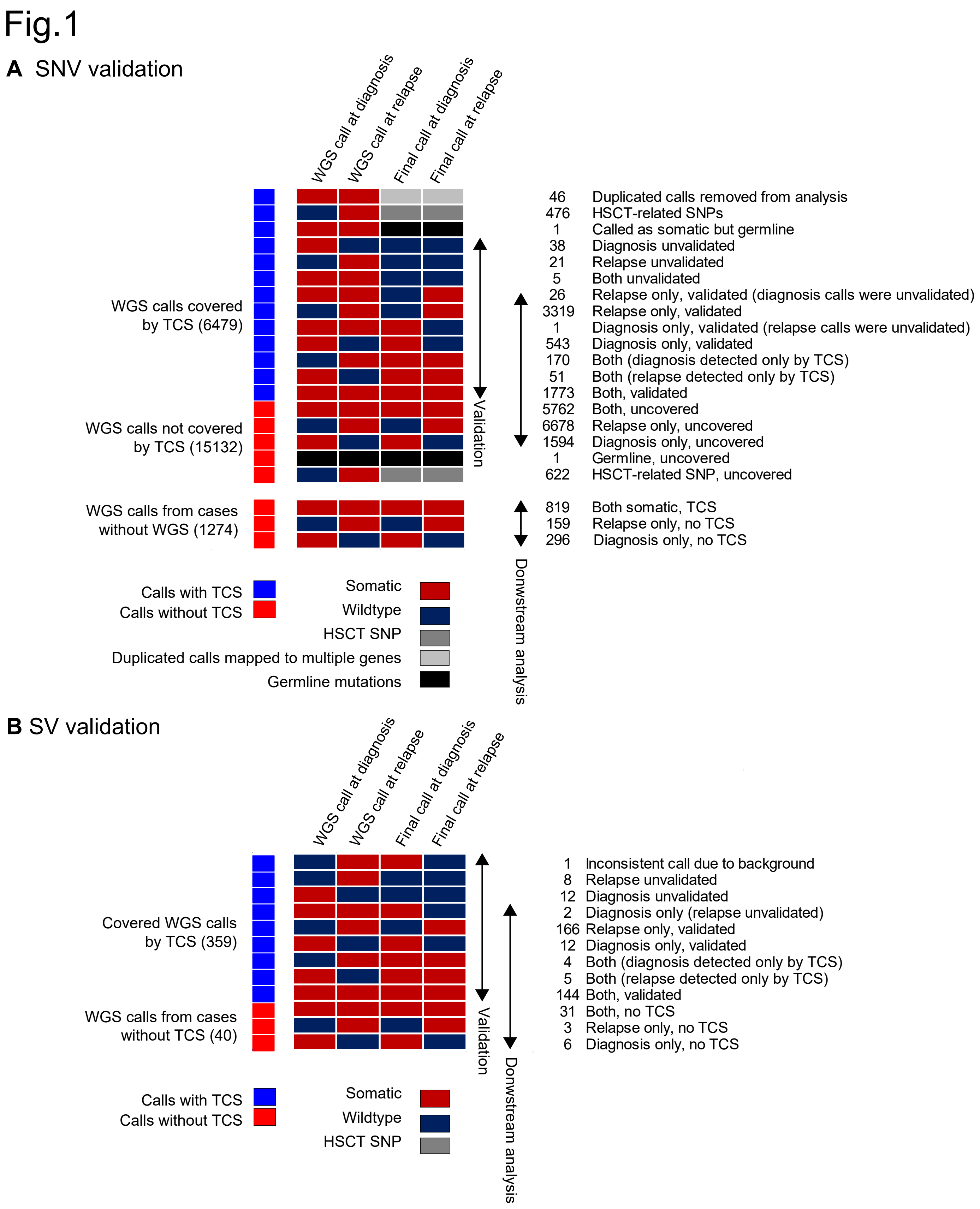


**Supplementary Figure 1. Validation of somatic alterations.** Validation status of single-nucleotide variants (SNVs) and insertions and deletions (indels, A) and structural variants (SV, B) found in this study. Each row represents calls from WGS and the final call at diagnosis and relapse. Each row represents validation status annotated on the right and the number of alterations. Abbreviations. WGS: whole-genome sequencing, TCS: targeted-capture sequencing, HSCT: hematopoietic stem cell transplant, SNP: single-nucleotide polymorphism.
