## Supplementary material for "Genetic Evolution of Pediatric Acute Myeloid Leukemia and Its Contribution to Disease Relapse": Fig S2

### Fig.2

#### A Diagnosis-Relapse pairs

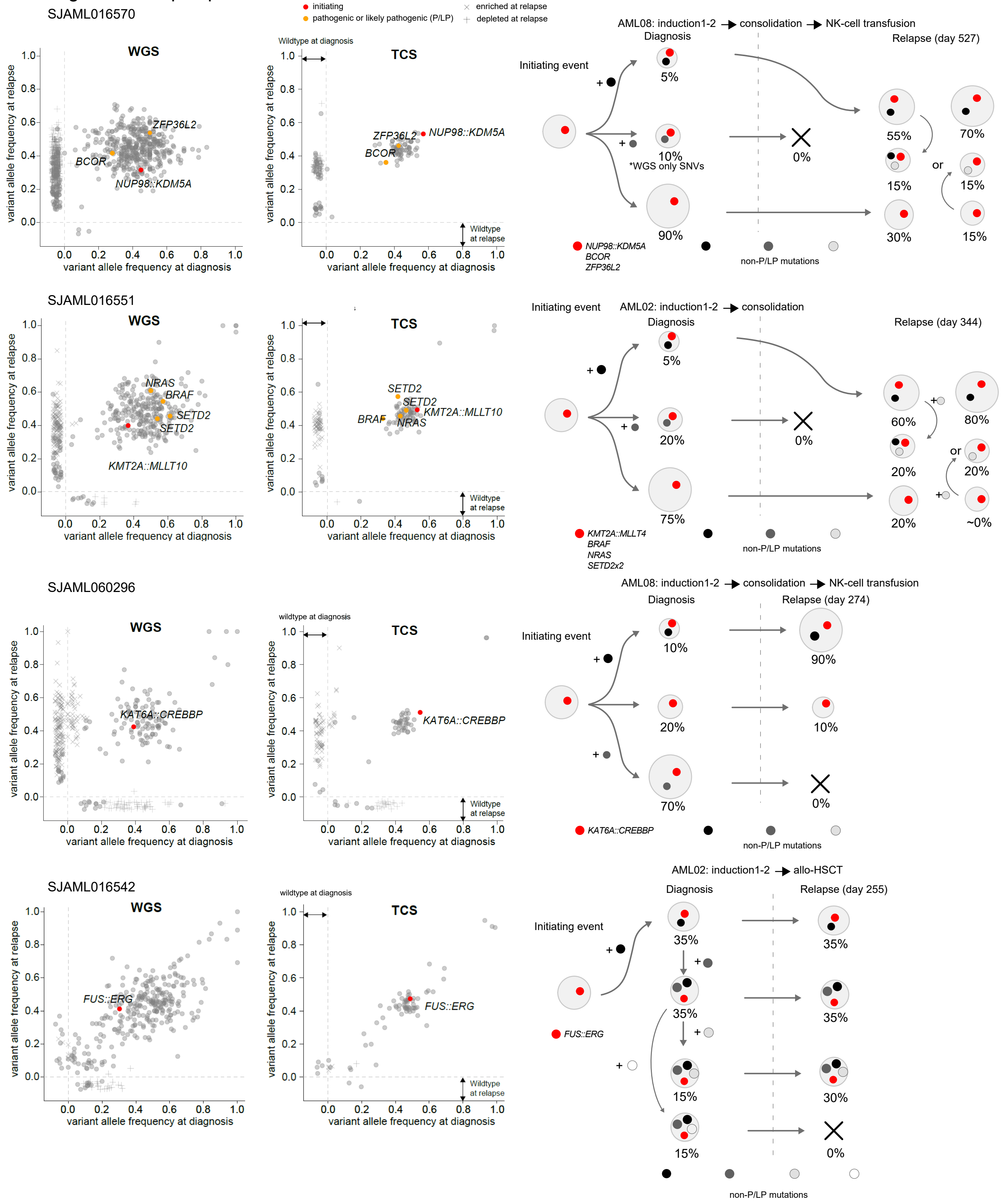

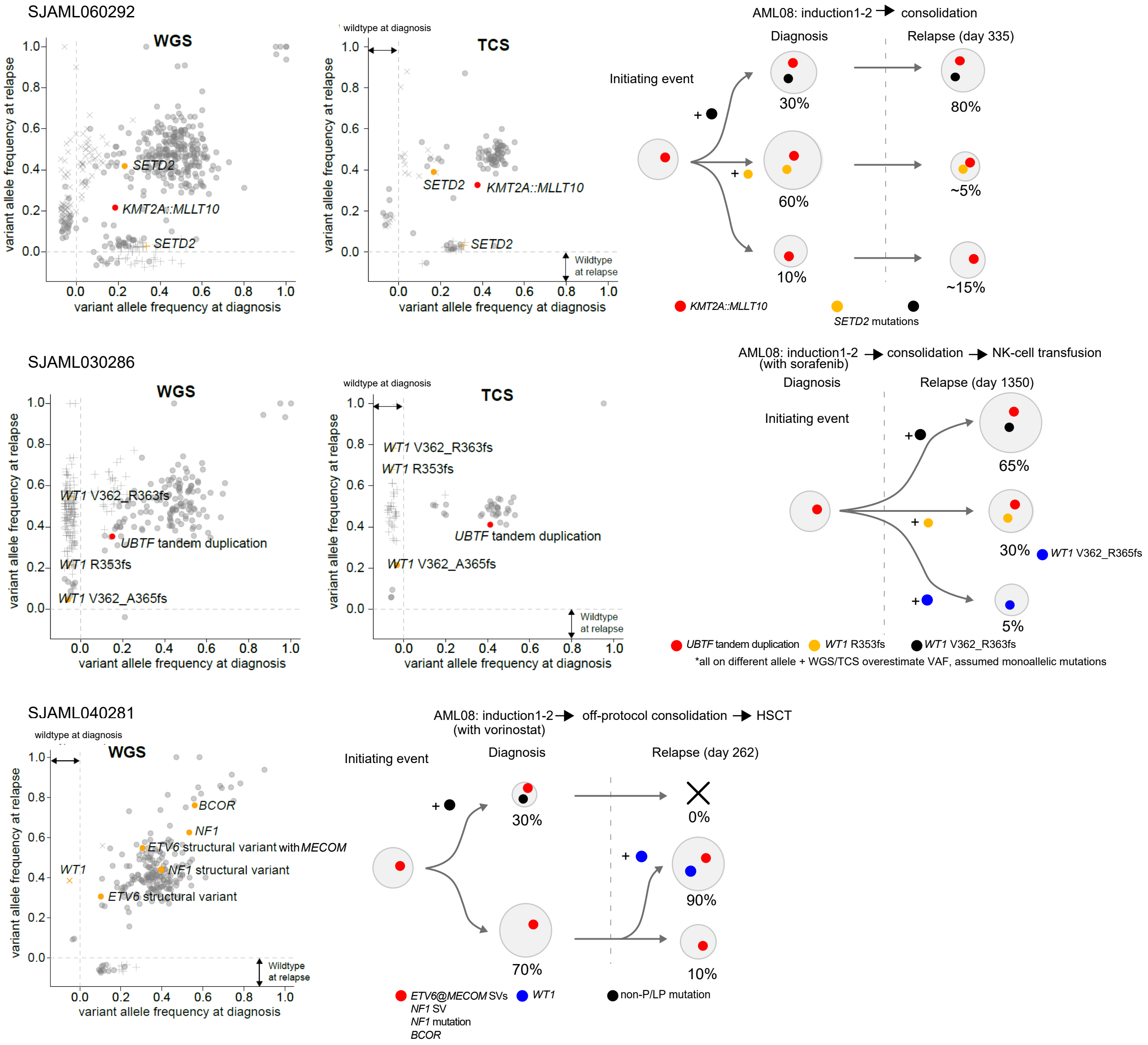

SJAML060292
WGS
TCS
variant allele frequency at relapse
variant allele frequency at diagnosis
wildtype at diagnosis
Wildtype at relapse
SETD2
KMT2A::MLLT10
SETD2
AML08: induction1-2 → consolidation
Diagnosis
Relapse (day 335)
Initiating event
30%
60%
10%
80%
~5%
~15%
KMT2A::MLLT10
SETD2 mutations
SJAML030286
WT1 V362_R363fs
UBTF tandem duplication
WT1 R353fs
WT1 V362_A365fs
AML08: induction1-2 (with sorafenib) → consolidation → NK-cell transfusion
Diagnosis
Relapse (day 1350)
Initiating event
65%
30%
5%
WT1 V362_R365fs
UBTF tandem duplication
WT1 R353fs
WT1 V362_R363fs
*all on different allele + WGS/TCS overestimate VAF, assumed monoallelic mutations
SJAML040281
BCOR
NF1
ETV6 structural variant with MECOM
WT1
NF1 structural variant
ETV6 structural variant
AML08: induction1-2 (with vorinostat) → off-protocol consolidation → HSCT
Initiating event
Diagnosis
Relapse (day 262)
30%
70%
0%
90%
10%
ETV6@MECOM SVs
NF1 SV
NF1 mutation
BCOR
WT1
non-P/LP mutation

**B** Relapse1-Relapse2 pairs

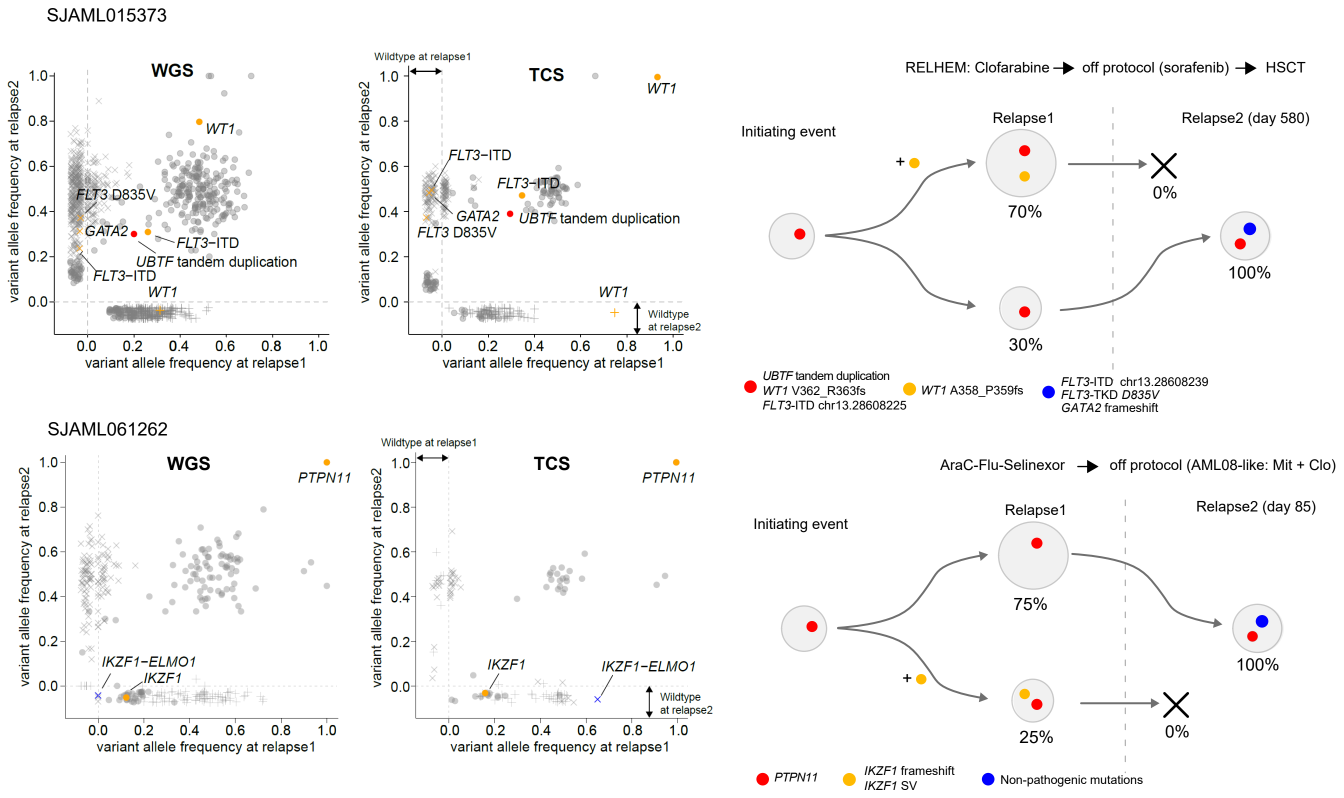


**Supplementary Figure 2. Clonal evolution patterns of representative cases from the cohort.** A. Diagnosis-relapse pairs (n=7) and B. Relapse-relapse pairs (n=2). On 2D plots of variant allele frequencies (VAFs), the colors of dots show the pathogenicity of somatic alterations, and the shapes of dots show enrichment at each time point. Undetected calls are shown away from the axes. On the schematics of clonal patterns, colored dots show mutations marking clones.
