## Supplementary material for "Genetic Evolution of Pediatric Acute Myeloid Leukemia and Its Contribution to Disease Relapse": Fig S3

### Fig.3

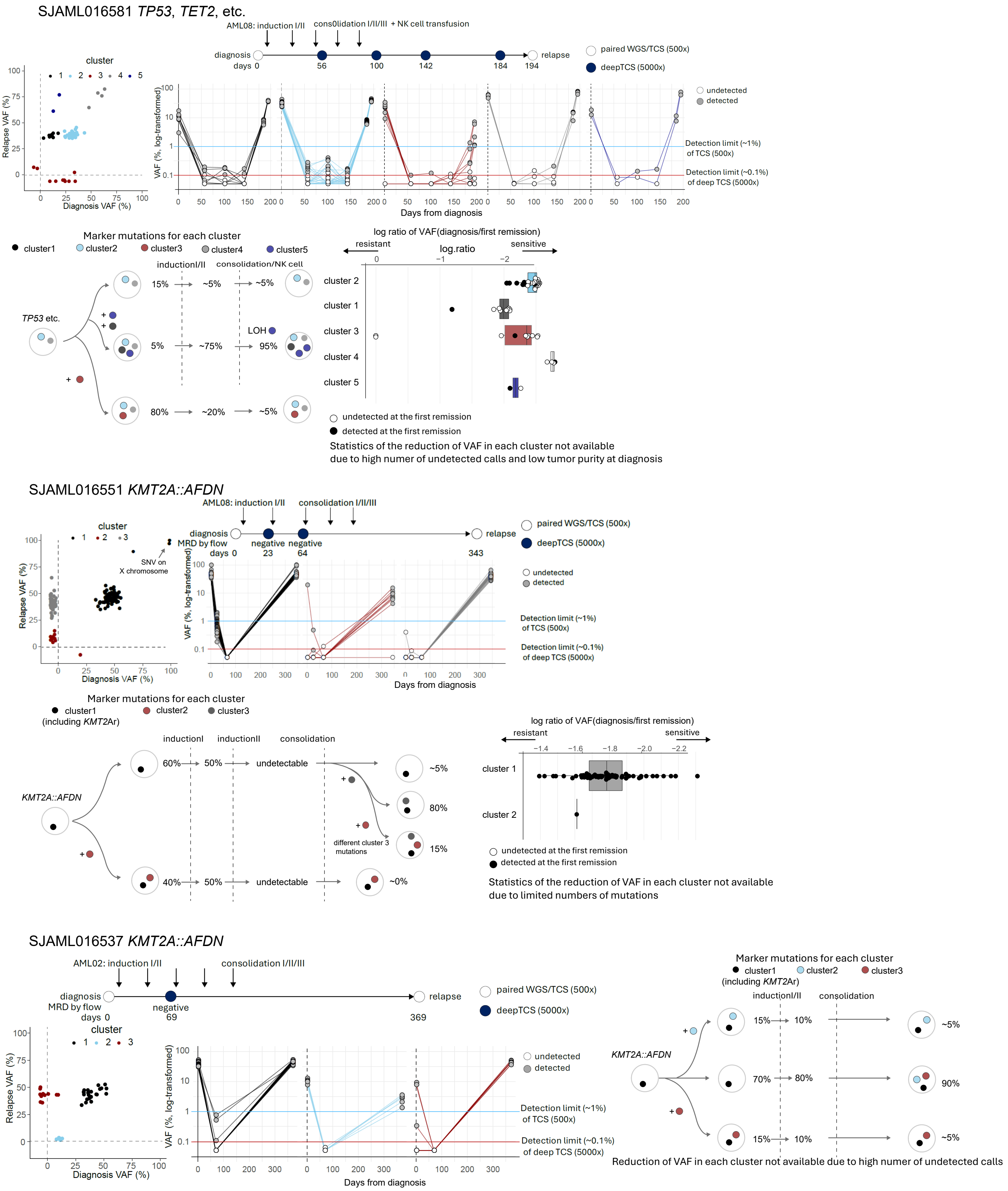

SJAML016581 TP53, TET2, etc.
AML08: induction I/II
consOlidation I/II/III + NK cell transfusion
diagnosis
days 0
56
100
142
184
194
relapse
paired WGS/TCS (500x)
deepTCS (5000x)
cluster
1 2 3 4 5
Relapse VAF (%)
Diagnosis VAF (%)
VAF (%, log-transformed)
undetected
detected
Detection limit (~1%) of TCS (500x)
Detection limit (~0.1%) of deep TCS (5000x)
Days from diagnosis
Marker mutations for each cluster
cluster1 cluster2 cluster3 cluster4 cluster5
inductionI/II
consolidation/NK cell
TP53 etc.
15% ~5% ~5%
5% ~75% 95%
LOH
80% ~20% ~5%
log ratio of VAF(diagnosis/first remission)
resistant
log.ratio
sensitive
cluster 2
cluster 1
cluster 3
cluster 4
cluster 5
undetected at the first remission
detected at the first remission
Statistics of the reduction of VAF in each cluster not available
due to high numer of undetected calls and low tumor purity at diagnosis
SJAML016551 KMT2A::AFDN
AML08: induction I/II
consolidation I/II/III
diagnosis
MRD by flow
days 0
negative
23
negative
64
343
relapse
paired WGS/TCS (500x)
deepTCS (5000x)
cluster
1 2 3
SNV on X chromosome
Relapse VAF (%)
Diagnosis VAF (%)
VAF (%, log-transformed)
undetected
detected
Detection limit (~1%) of TCS (500x)
Detection limit (~0.1%) of deep TCS (5000x)
Days from diagnosis
Marker mutations for each cluster
cluster1 (including KMT2Ar)
cluster2
cluster3
inductionI
inductionII
consolidation
KMT2A::AFDN
60% 50% undetectable ~5%
80%
different cluster 3 mutations
15%
40% 50% undetectable ~0%
log ratio of VAF(diagnosis/first remission)
resistant
sensitive
cluster 1
cluster 2
undetected at the first remission
detected at the first remission
Statistics of the reduction of VAF in each cluster not available
due to limited numbers of mutations
SJAML016537 KMT2A::AFDN
AML02: induction I/II
consolidation I/II/III
diagnosis
MRD by flow
days 0
negative
69
369
relapse
paired WGS/TCS (500x)
deepTCS (5000x)
cluster
1 2 3
Relapse VAF (%)
Diagnosis VAF (%)
VAF (%, log-transformed)
undetected
detected
Detection limit (~1%) of TCS (500x)
Detection limit (~0.1%) of deep TCS (5000x)
Days from diagnosis
Marker mutations for each cluster
cluster1 (including KMT2Ar)
cluster2
cluster3
inductionI/II
consolidation
KMT2A::AFDN
15% 10% ~5%
70% 80% 90%
15% 10% ~5%
Reduction of VAF in each cluster not available due to high numer of undetected calls

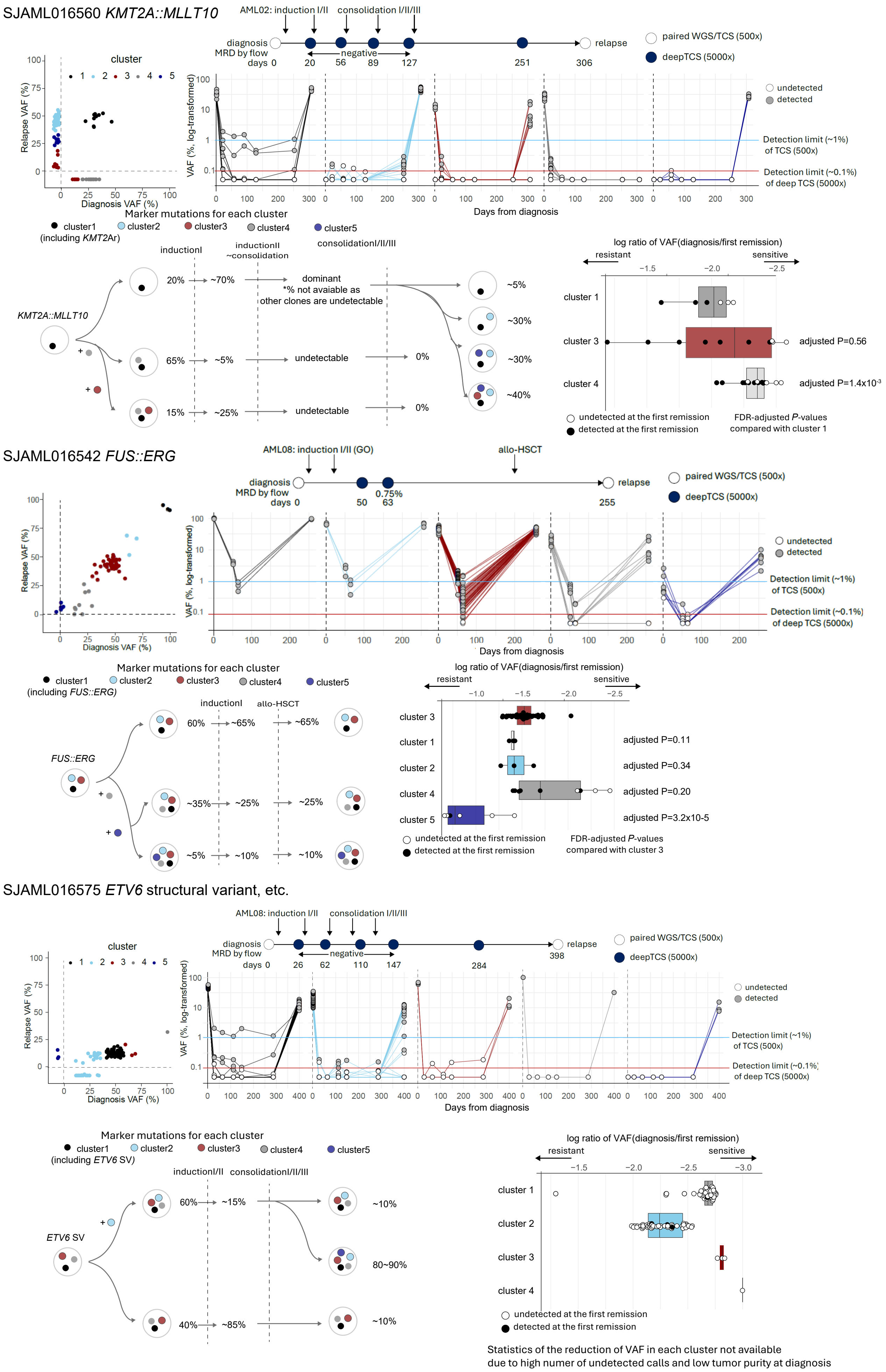
SJAML016560 KMT2A::MLLT10
AML02: induction I/II
consolidation I/II/III
diagnosis
MRD by flow
days 0
negative
20
56
89
127
251
306
relapse
paired WGS/TCS (500x)
deepTCS (5000x)
cluster
1 2 3 4 5
Relapse VAF (%)
Diagnosis VAF (%)
VAF (%, log-transformed)
Days from diagnosis
undetected
detected
Detection limit (~1%) of TCS (500x)
Detection limit (~0.1%) of deep TCS (5000x)
Marker mutations for each cluster
cluster1 (including KMT2Ar)
cluster2
cluster3
cluster4
cluster5
inductionI
inductionII ~consolidation
consolidationI/II/III
KMT2A::MLLT10
20%
~70%
dominant
*% not avaiable as other clones are undetectable
65%
~5%
undetectable
0%
15%
~25%
undetectable
0%
~5%
~30%
~30%
~40%
log ratio of VAF(diagnosis/first remission)
resistant
sensitive
cluster 1
cluster 3
cluster 4
adjusted P=0.56
adjusted P=1.4x10-3
undetected at the first remission
detected at the first remission
FDR-adjusted P-values compared with cluster 1
SJAML016542 FUS::ERG
AML08: induction I/II (GO)
allo-HSCT
diagnosis
MRD by flow
days 0
50
0.75%
63
255
relapse
paired WGS/TCS (500x)
deepTCS (5000x)
Relapse VAF (%)
Diagnosis VAF (%)
VAF (%, log-transformed)
Days from diagnosis
undetected
detected
Detection limit (~1%) of TCS (500x)
Detection limit (~0.1%) of deep TCS (5000x)
Marker mutations for each cluster
cluster1 (including FUS::ERG)
cluster2
cluster3
cluster4
cluster5
inductionI
allo-HSCT
FUS::ERG
60%
~65%
~65%
~35%
~25%
~25%
~5%
~10%
~10%
log ratio of VAF(diagnosis/first remission)
resistant
sensitive
cluster 3
cluster 1
cluster 2
cluster 4
cluster 5
adjusted P=0.11
adjusted P=0.34
adjusted P=0.20
adjusted P=3.2x10-5
undetected at the first remission
detected at the first remission
FDR-adjusted P-values compared with cluster 3
SJAML016575 ETV6 structural variant, etc.
AML08: induction I/II
consolidation I/II/III
diagnosis
MRD by flow
days 0
negative
26
62
110
147
284
relapse
398
paired WGS/TCS (500x)
deepTCS (5000x)
cluster
1 2 3 4 5
Relapse VAF (%)
Diagnosis VAF (%)
VAF (%, log-transformed)
Days from diagnosis
undetected
detected
Detection limit (~1%) of TCS (500x)
Detection limit (~0.1%) of deep TCS (5000x)
Marker mutations for each cluster
cluster1 (including ETV6 SV)
cluster2
cluster3
cluster4
cluster5
inductionI/II
consolidationI/II/III
ETV6 SV
60%
~15%
~10%
80~90%
40%
~85%
~10%
log ratio of VAF(diagnosis/first remission)
resistant
sensitive
cluster 1
cluster 2
cluster 3
cluster 4
undetected at the first remission
detected at the first remission
Statistics of the reduction of VAF in each cluster not available
due to high numer of undetected calls and low tumor purity at diagnosis

**Supplementary Figure 3. Serial sequencing of remission samples in six cases.** Mutational patterns at diagnosis and relapse, clinical course, changes in VAFs along time points, inferred clonal structures, and comparison of log10 fold changes of VAFs at diagnosis and immediately after induction therapy of six cases not in Figure 5. For plots and fold changes of calls with no mutant read count, a VAF of 0.0005 (half of the expected limit of detection of deep sequencing with 5000x coverage) was used as a value. Statistical significance of the reduction of VAFs was assessed by the Wilcoxon rank-sum test, followed by adjustment by the Benjamini-Hochberg procedure, whereas plots or statistical tests for cases with insufficient somatic calls were excluded. Lines of the box plots represent the 25% quantile, median, and 75% quantile. The upper and lower whisker represents the higher value of maxima or 1.5 x IQR and the lower value of minima or 1.5 x IQR, respectively.
