## Supplementary material for "Genetic Evolution of Pediatric Acute Myeloid Leukemia and Its Contribution to Disease Relapse": Fig S4

### Supplementary Figure 4

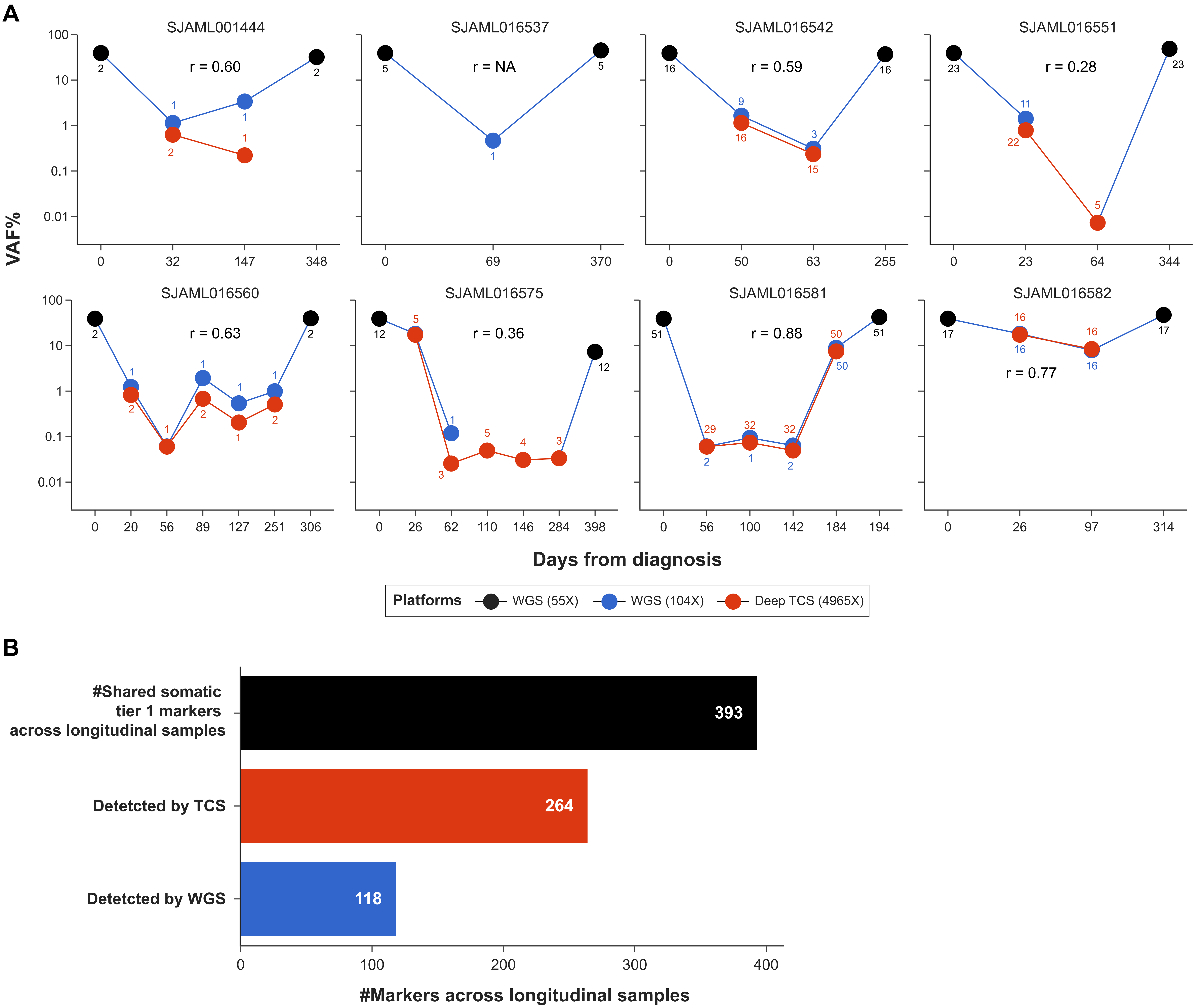


**Supplementary Figure 4. Detetction of shared somatic tier 1/coding SNV/Indels by higher-depth WGS and deep TCS.**
**A.** Shown are the patient-wise cumulative variant allele frequencies (VAF, Y-axis) of tier 1/coding somatic markers detected in shared/retrospective manner across longitudinal samples for eight AML patients using whole genome at 100X, compared with deep TCS at 5000X.
The days of sample collection from diagnosis are shown on X-axis. The points shown in black represent cumulative VAFs of makers detected in diagnosis/relapse timepoint samples sequenced using standard WGS (median depth: 55X), those in blue represent the same for longitudinal samples sequenced using relatively high depth WGS (median depth: 104X) and those in red represent longitudinal samples with deep TCS data (median depth: 4965X). The number of markers detected are shown along points and were used to generate cumulative VAFs from respective sequencing platforms at given sampling points. The patient-wise Pearson's correlation coefficients (R) between VAFs at valid loci (see Methods) captured by high depth WGS and deep sequencing platforms over longitudinal timepoints are shown.
**B.** Number of shared tier1/coding somatic SNV/Indels detetcted by higher-depth WGS and deep TCS paltforms. Bar in black shows the total number of shared markers across longitudinal timepoints (#observations) whereas those in red and blue respectively indicate detetcted markers by deep TCS and higher-depth WGS.

WGS: whole genome sequencing; TCS: targeted capture sequencing
