## Supplementary material for "Genetic Evolution of Pediatric Acute Myeloid Leukemia and Its Contribution to Disease Relapse": Fig S5

### Supplementary Figure 5

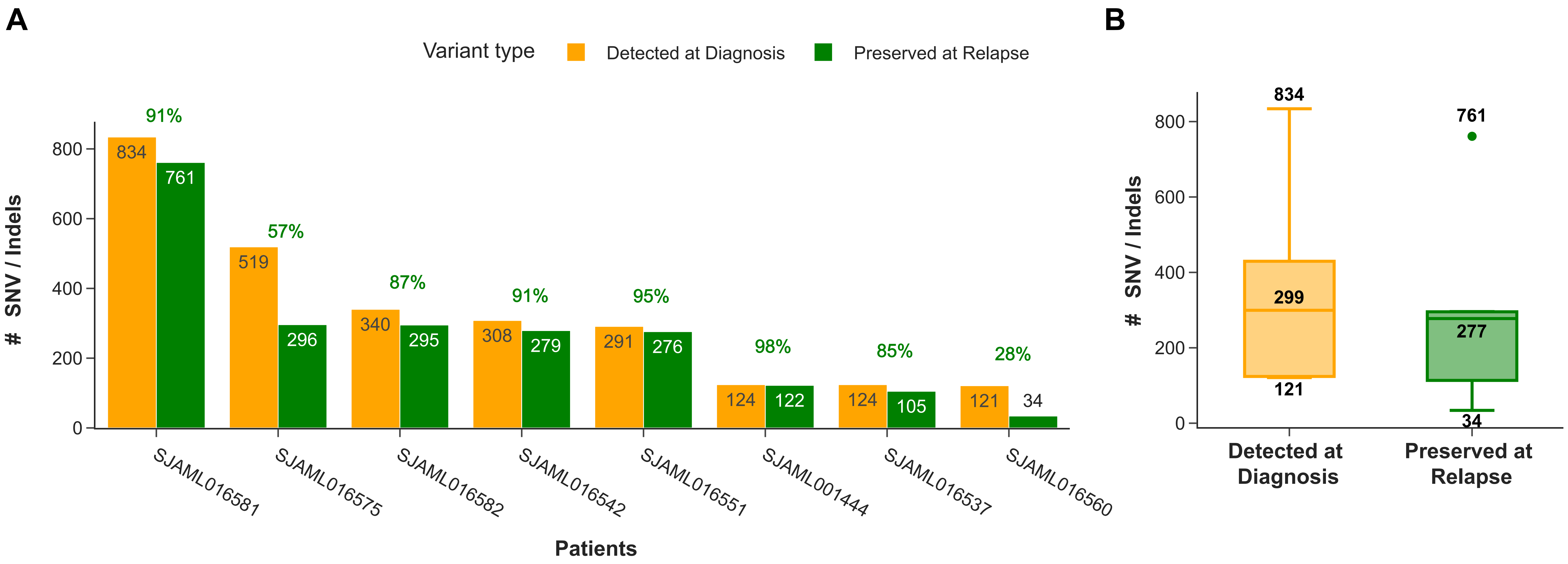


**Supplementary Figure 5. Number of SNV/Indels detetcted at diagnosis and preserved till relapse.**
**A.** Orange bars show patient-wise number of SNV/Indels detetcted at diagnosis whereas green bars indicate those preserved at relapse. Percentage of preserved markers from diagnosis are shown on top of each bar pair per patient.
**B.** Box plots showing distribution of number of SNV/Indels detetcted at diagnosis (orange) and those preserved at relapse (green). Minimum, median and maximum number of markers are indicated along boxplots for each category.
